# Gene expression profiling beyond Breslow thickness and ulceration for prediction of distant metastases in early-stage melanoma: the population-based Dutch Early-Stage Melanoma (D-ESMEL) study

**DOI:** 10.1101/2025.11.10.25339906

**Authors:** Catherine Zhou, Yan-Ting Chen, Antien L. Mooyaart, Erik T. Valent, Lara Pozza, Daphne M.S. Huigh, Tamar E.C. Nijsten, Loes M. Hollestein

## Abstract

**Purpose:** Despite their central role in the current staging system, Breslow thickness and ulceration do not fully identify early-stage melanoma patients who will develop distant metastases. We assessed whether gene expression profiling (GEP) improves prediction of distant metastases beyond standard staging factors in early-stage melanoma.

**Methods:** Data were derived from the population-based Dutch Early-Stage Melanoma (D-ESMEL) study, including a matched discovery set of 442 stage I/II melanomas (221 case-control pairs) and a validation cohort of 308 melanomas nested within 5,815 patients. The discovery set was used to identify genes associated with distant metastases, independent of age, sex, Breslow thickness, and ulceration. The validation cohort was partitioned into model development and independent validation subsets. Candidate genes from the discovery set were used to develop and validate a GEP model, evaluated by weighted area under the curve (AUC) and concordance index (C-index).

**Results:** RNA sequencing succeeded for 356 melanomas in the discovery set, 200 in the model development subset, and 94 melanomas in the independent validation subset. Differential gene expression analyses and modeling identified 558 candidate genes. In the independent validation subset, the GEP model achieved a weighted AUC of 0.77 (95% CI, 0.66-0.86) and weighted C-index of 0.79 (95% CI, 0.69-0.88), comparable to the clinical model based on Breslow thickness and ulceration (weighted AUC 0.82 (95% CI, 0.73-0.90), weighted C-index 0.84 (95% CI, 0.76-0.91)). Integration of GEP with the clinical model did not improve accuracy. Gene set enrichment analyses showed enrichment of proliferative and stress-related pathways.

**Conclusion:** While GEP captured biologically relevant signals, its predictive accuracy for distant metastases was comparable to that of Breslow thickness and ulceration in a population-based early-stage melanoma cohort.

## Introduction

Accurate prognostication in early-stage cutaneous melanoma is critical for identifying patients at risk of progression. The American Joint Committee on Cancer (AJCC) staging system, based on Breslow thickness and ulceration, has demonstrated robust prognostic utility ^1^. Nevertheless, these clinicopathological parameters alone do not fully capture the biological heterogeneity that drives variability in patient outcomes, as a subset of early-stage patients still develops distant metastases despite favorable initial staging ^2–4^. Approximately 41% of melanoma deaths and 60% of distant metastases occur in patients initially diagnosed with stage I/II disease ^2,4^. This underscores the need for molecular biomarkers that extend beyond current staging parameters to better capture the drivers of progression in early-stage melanoma.

Gene expression profiling (GEP) has emerged as a promising approach to refine risk prediction by identifying molecular signatures ^5,6^. Several GEP assays have been developed for melanoma risk stratification ^7–11^. CP-GEP (commercially known as the Merlin assay, SkylineDx) was developed to identify patients at low risk of sentinel lymph node metastasis who may forgo sentinel lymph node biopsy (SLNB) and has been validated in multiple studies ^12–14^. Other assays, including DecisionDx-Melanoma (Castle Biosciences, Inc.), MelaGenix (NeraCare GmbH), LMC_150 (Leeds Melanoma Cohort) and Cam_121 (Cambridge Cancer Center), have demonstrated their ability to identify patient subgroups at higher risk of progression ^8–11^. Despite their potential to improve risk stratification, current clinical guidelines from the National Comprehensive Cancer Network (NCCN), the Society of Surgical Oncology (SSO) and the American Academy of Dermatology (AAD) do not yet recommend routine GEP testing, awaiting results from prospective validation in large datasets of unselected patients ^15–17^.

Development and validation of clinically useful GEP models require large, well-powered studies with sufficient metastatic events to ensure stable, generalizable predictions and to demonstrate clinical utility ^5,6^. A critical concern is insufficient adjustment for established prognostic variables, which may result in high-risk GEP classifications that primarily reflect known high-risk features rather than providing independent prognostic information ^18^. While multivariable adjustment can address this issue, application to cohorts with limited events risks statistical instability, model overfitting, and wide confidence intervals despite high hazard ratios, as illustrated in prior evaluations ^9,19^.

The Dutch Early-Stage Melanoma (D-ESMEL) study was designed to address these challenges by evaluating the value of GEP for identifying early-stage melanoma patients at risk of distant metastasis beyond well-established clinicopathological factors. We employed a matched case-control discovery set to control for key prognostic variables, followed by model development and independent validation in a nationwide cohort to enable robust performance assessment ^20^. Using this study design, we sought to develop and validate a GEP model for predicting distant metastasis risk in early-stage melanoma and determine whether it provides added value over current clinicopathological staging factors.

## Methods

### Study design and patient population

This study utilized the methodological framework of the D-ESMEL study, drawing data from the Netherlands Cancer Registry (NCR) and the Dutch Nationwide Pathology Databank (PALGA), as previously described ^20^. In brief, the discovery set comprised 221 matched case-control pairs of stage I/II melanoma patients matched on age, sex, Breslow thickness, and ulceration. Cases were patients who developed distant metastases (stage IV disease based on the AJCC); controls were patients who remained distant metastases-free within the time period in which the matched case developed distant metastases. This design enabled identification of candidate genes independent of known clinicopathological factors.

To develop and validate a prognostic model, we established a validation cohort using a nested case-control (NCC) design within a nationwide cohort of 5,815 patients diagnosed with stage I/II melanoma between 2016 and 2020. This included 154 case-control sets matched on follow-up time. The validation cohort was split into two-thirds for model development and one-third for independent validation. The NCC design permits estimation of absolute risks ^21–23^.

This study was approved by the Erasmus MC Ethics Committee (MEC-2018-1738), the scientific committee of the NCR and the Privacy Committee of PALGA. The code of conduct for responsible use of human residual tissue by the Federation of Dutch Medical Scientific Societies was followed ^24^.

RNA extraction, library preparation, sequencing, and bioinformatic processing are described in detail in Supplementary Methods. The number of samples yielding successful transcriptomic data is reported in the Results section.

### Differential gene expression and modelling in the discovery set

To identify candidate genes predictive of distant metastasis, we combined differential gene expression analysis (DGEA) with penalized discriminative modeling in the discovery set. DGEA was conducted using multiple statistical frameworks, and the top-ranked genes were aggregated. In parallel, least absolute shrinkage and selection operator (LASSO)-penalized conditional logistic regression approach with double-loop cross-validation (DLCV) was used to select and internally validate predictive genes ^25^.

Candidate genes were selected as those with significantly higher selection frequency, assessed through permutation testing. Detailed methodological procedures are provided in Supplementary Figure 1 and Supplementary Methods.

### Model development

Following identification of candidate genes in the discovery set, model development was conducted in two-thirds of the validation cohort (hereafter referred to as the model development subset) to assess generalizability and predictive accuracy of the gene set.

First, we performed DGEA using DESeq2 ^26^ and limma ^27^ to determine how many genes identified in the discovery set remained differentially expressed in the model development subset. Models were conditioned for the matching variables from the discovery set and AJCC stage.

To assess the predictive performance of the gene set identified through DGEA and DLCV in the discovery set, we constructed baseline models using logistic regression with LASSO regularization: (1) clinicopathological variables (Breslow thickness and ulceration), (2) AJCC stage, (3) matching variables, (4) gene expression data, and (5) an integrated model combining clinical variables and gene expression.

All genes identified in the discovery set were initially included as input features. Alternative feature selection strategies were explored, including adaptive LASSO and ridge regression. Additionally, we examined whether subsets of 558 genes with higher selection frequency across models improved predictive accuracy.

Discriminative performance was assessed using both weighted area under the curve (AUC) and weighted concordance index (C-index) with corresponding 95% confidence intervals (95% CI). Given the NCC design and complete follow-up within the nationwide source cohort, sampling weights were applied to generate representative performance estimates ^21^. These weights were based on the distribution of follow-up time, as controls were randomly sampled without matching on prognostic factors. Internal validation consisted of 200 bootstrap iterations, where matched case-control pairs were kept together during resampling. The .632+ estimator was used for internally validated metrics ^28^.

### Independent validation

The best-performing and most robust models from the model development subset were applied to the remaining one-third of the validation cohort (hereafter referred to as the independent validation subset), which was withheld during model development, to evaluate their predictive performance.

Weighted AUC and C-index with corresponding 95% CI were recalculated using the methods previously described. Sampling weights were applied based on the corresponding one-third of the nationwide source cohort. This held-out subset enabled assessment of model stability, potential overfitting, and reproducibility.

### GEP models trained within the model development subset

In a complementary analysis, we trained GEP models entirely within the model development subset, without relying on genes identified in the discovery set. Instead, models were built using all genes with reliable expression measurements, defined as log_2_(TPM +1) > 0.5 in at least half of the model development subset and limited to protein-coding and long non-coding RNA (*n* = 16,661 genes). Models were trained using 200 bootstrap iterations with LASSO regularization across input sets of 50 to 1000 genes with the highest variances. Performance was assessed using weighted AUC and weighted C-index in the model development subset and the independent validation subset.

### Gene set enrichment analyses

To investigate the biological pathways associated with metastatic progression and established prognostic factors, we performed gene set enrichment analysis using the MSigDB Hallmark gene sets ^29^ and the DGEA on the model development subset of the validation cohort. Three complementary analyses were performed: (1) cases versus controls conditioned on matching variables, (2) melanomas with Breslow thickness >2 mm versus ≤2 mm, and (3) melanomas with versus without ulceration. Genes were ranked by moderated t-statistics from limma, and enrichment scores were calculated using the fgsea algorithm. Significance of enrichment was assessed based on normalized enrichment scores (NES) and adjusted p-values (false discovery rate <0.05).

### Exploratory analyses of publicly available gene sets

We conducted exploratory analyses using previously published gene sets as positive methodological controls ^7–11^. Although these gene signatures differ in endpoints, technical platforms, and intended patient populations compared with our study, we hypothesized that they would still show association with distant metastasis in our cohort, thereby serving as an internal validation of our methodology. We examined the distribution of differentially expressed genes from each signature using volcano plots.

Each gene set was then used as input for logistic regression models with LASSO regularization and performance evaluated using weighted AUC and weighted C-index metrics with corresponding 95% CI in the model development subset. Accordingly, our analyses should be interpreted as exploratory checks of methodological consistency, not as comparative assessments of clinical performance.

## Results

### Discovery set and validation cohort

The characteristics of the discovery set, the validation cohort, including the model development and the independent validation subsets, and their respective source cohorts have been described previously ^20^. For the present analysis, only samples with successful RNA sequencing (RNAseq) after quality control measures were included (Supplementary Figure 2). In total, RNAseq data were successfully generated for 178 case-control pairs in the discovery set, comprising 356 patients matched on age, sex, Breslow thickness, and ulceration. In the validation cohort, RNAseq data were successfully generated for 147 case-control pairs. This cohort was divided into two subsets: the model development subset (*n* = 100 pairs) and the independent validation subset (*n* = 47 pairs). The corresponding source cohort sizes (3,776 and 1,775 patients, respectively) were calculated based on the sampling fractions of these subsets within the same nationwide cohort (n = 5,551) (Table 1).

**Table 1.**
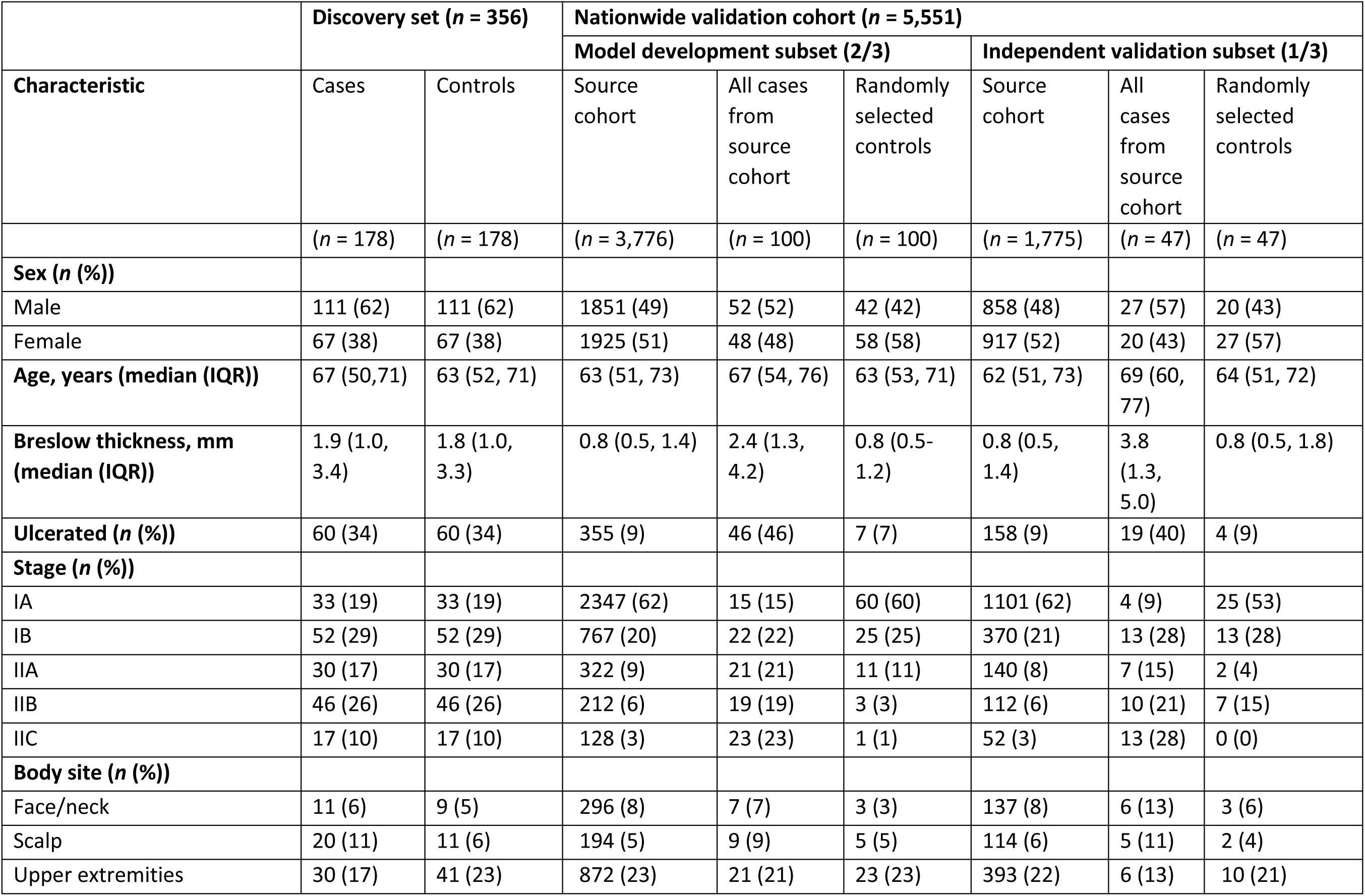
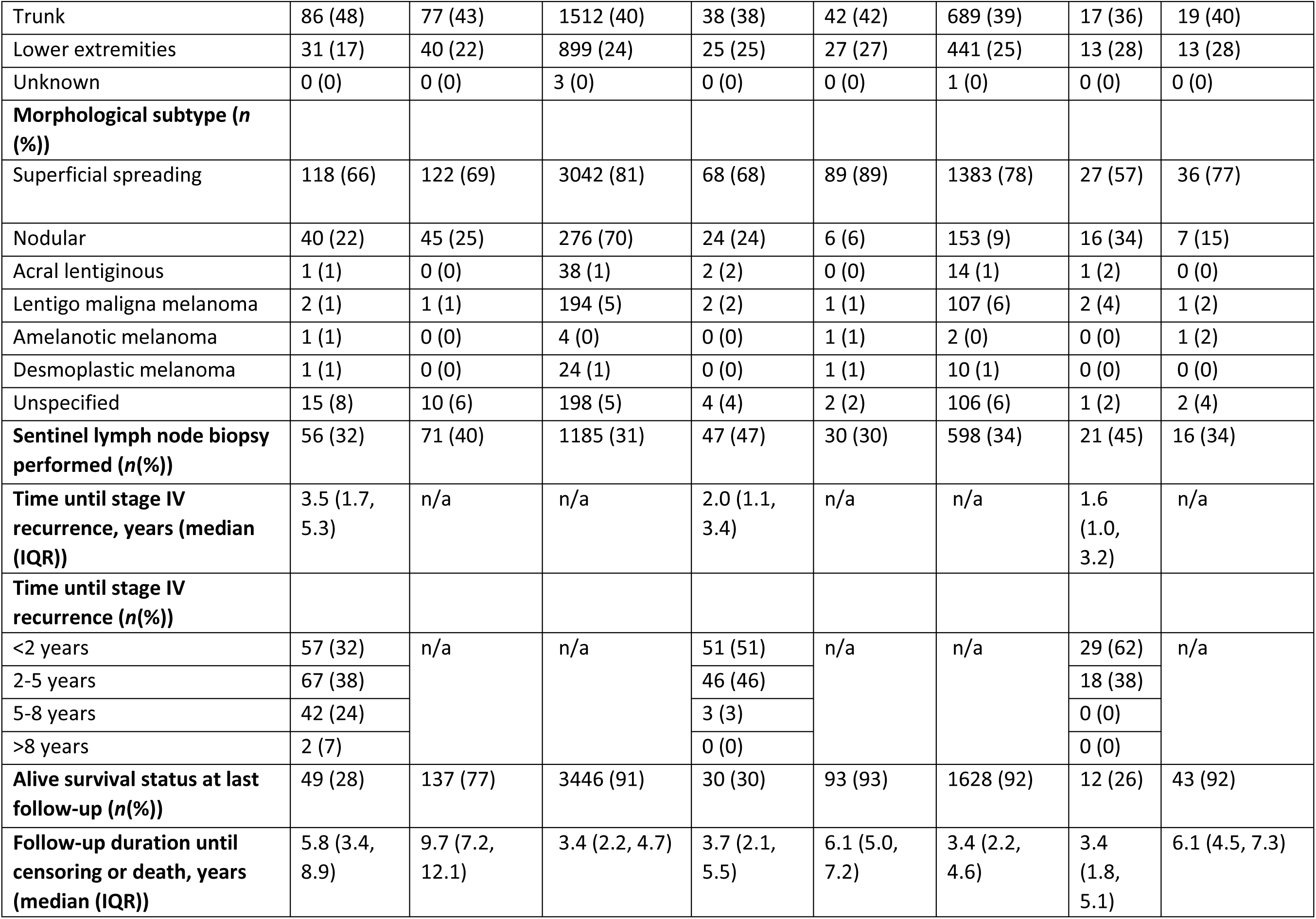
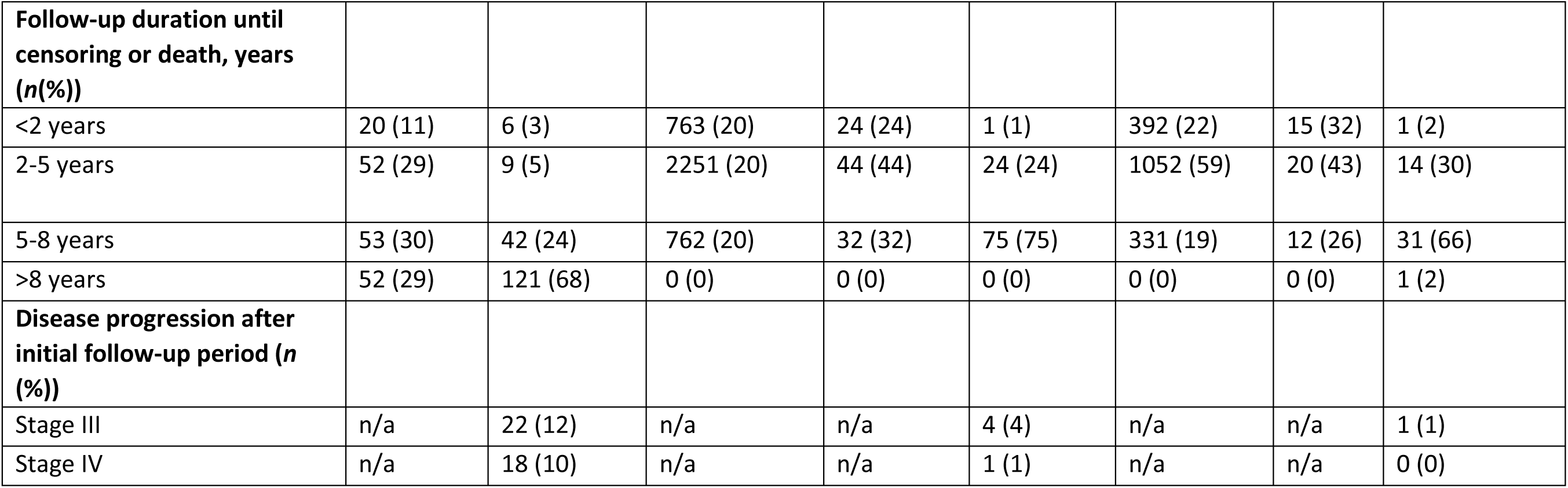
Clinical characteristics of the discovery set, validation cohorts and its nationwide source cohort. The validation cohort was randomly split into two-thirds for model development and one-third for independent model validation.

### Candidate prognostic genes based on the discovery set

We identified 288 differentially expressed genes between cases and controls using a union-based selection approach across multiple statistical models. The selection was based on the top 100 genes with the smallest *p*-values from seven differential expression models, each conditioned on either matching variables or the paired nature of the data. We ultimately did not use DESeq2 conditioned on pairs due to unstable dispersion estimates. The resulting volcano plots for all models are shown in Figure 1.

**Fig 1.**
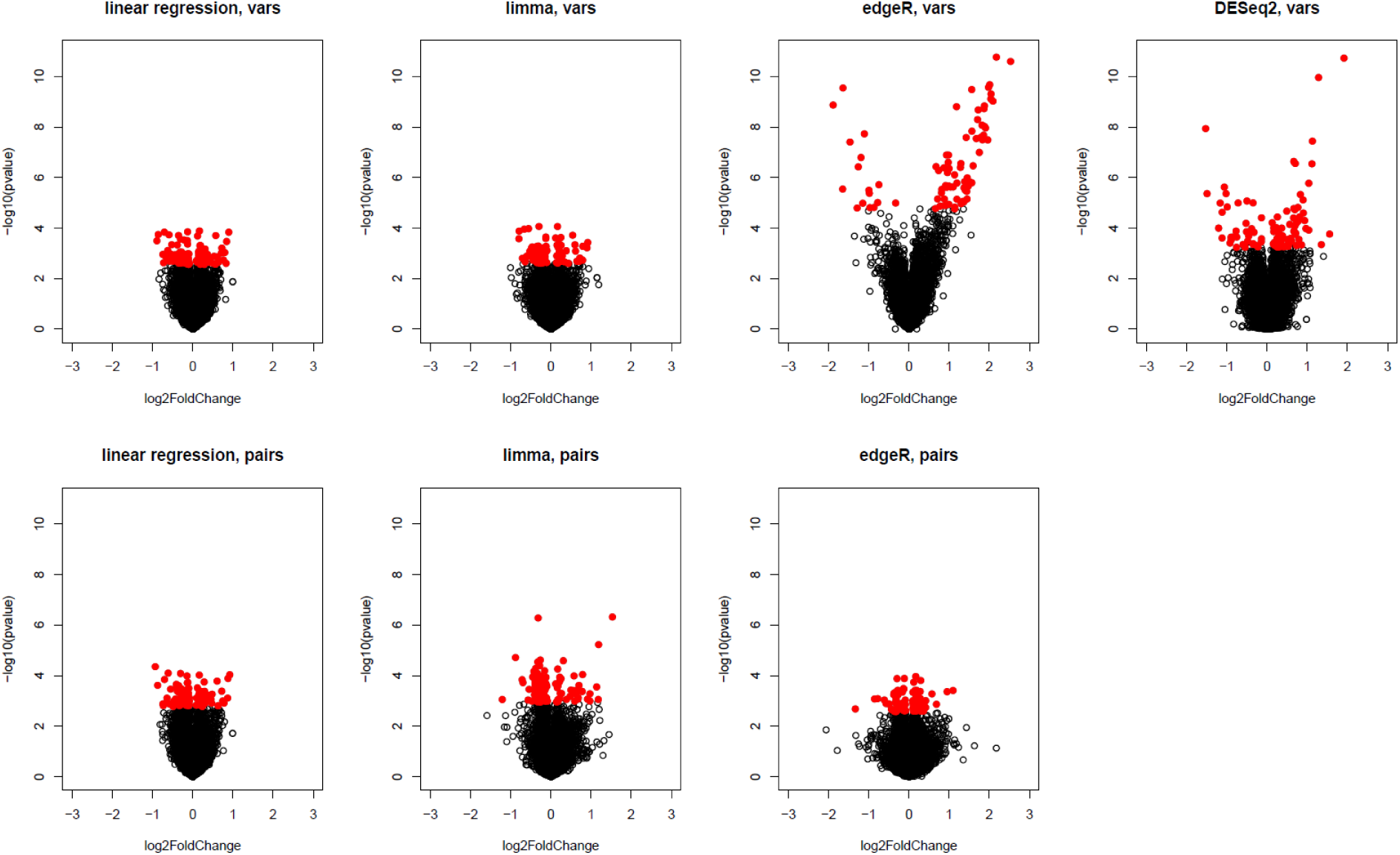
Volcano plots from seven differential gene expression analysis models comparing cases and matched controls in the discovery set. Each model was conditioned on matching variables (top) and on case-control pairs (bottom). Genes highlighted in red represent the 100 genes with the smallest p-values in each model.

LASSO conditional logistic regression with DLCV resulted in 350 genes that were most frequently (defined as ≥ 20%) selected. Permutation testing with 100 randomized case-control label assignments (Supplementary Figure 1) demonstrated that models built on original data performed better than the models based on permuted data, with original data models achieving an AUC of ∼0.6 with 500 or more input genes (Supplementary Figure 3). Combining both approaches, we constructed a final gene set comprising 558 unique genes, including 80 overlapping entries. A list of these genes is provided in Supplementary Table 1.

### Model development

DGEA in the model development subset without adjustment revealed 8,001 significantly differentially expressed genes. When conditioning on matching variables of the discovery set, the number of significantly differentially expressed genes was markedly reduced (*n* = 526). No significantly differentially expressed genes remained when conditioning on AJCC stage (Supplementary Figure 4A-C). The 558 genes identified in the discovery set were not more significant than other genes in the model development subset, when conditioned for matching variables (Supplementary Figure 4D and E).

Next, we compared the predictive performance of clinical, gene-based, and integrated models. Breslow thickness combined with ulceration achieved a weighted AUC of 0.82 (95% CI, 0.78-0.86) and a weighted C-index of 0.85 (95% CI, 0.72-0.88), which was comparable to models based on AJCC stage (weighted AUC 0.79 [95% CI, 0.75-0.82], weighted C-index 0.83 [95% CI, 0.79-0.86]) or on the full set of matching variables from the discovery set (age, sex, ulceration, and Breslow thickness) (weighted AUC 0.79, [95% CI, 0.74-0.83], weighted C-index 0.83, [95% CI, 0.78-0.86]). Adding the 558 genes to Breslow thickness and ulceration did not improve performance (weighted AUC 0.80 [95% CI, 0.74-0.88], weighted C-index 0.83 [95% CI, 0.77-0.89]) compared to Breslow thickness and ulceration alone (Figure 2A; Table 2).

**Fig 2.**
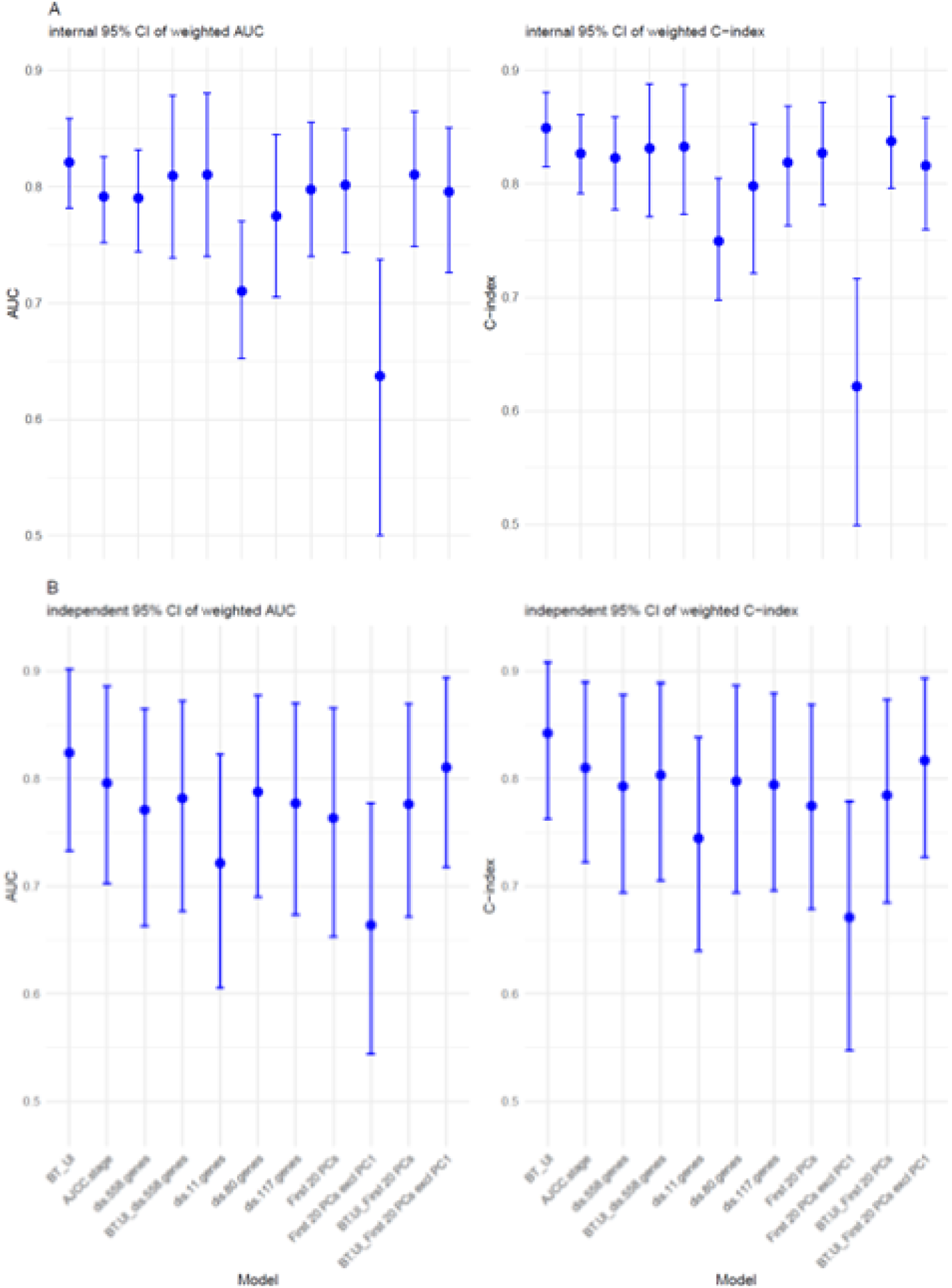
Weighted area under the curve (AUC) (left) and C-index (right) for all prediction models, evaluated in the model development subset (A) and independent validation subset (B). Models include: Breslow thickness plus ulceration (BT_Ul); American Joint Committee of Cancer (AJCC) stage; 558-gene signature; 558-gene signature plus BT_Ul; 11-gene, 80-gene, and 117-gene subsets; first 20 principal components (PC20); PC20 excluding PC1; PC20 plus BT_Ul; and PC20 excluding PC1 plus BT_Ul.

**Table 2.** Performance of prognostic models during model development and independent validation. Weighted AUC and C-index are shown for each model, using 0.632+ correction in the model development subset.*variable was included as continuous variable. AJCC = American Joint Committee of Cancer. GEP = gene expression profile.

| Model | Features | Model development subset |  | Independent validation subset |  |
| --- | --- | --- | --- | --- | --- |
|  |  | Weighted AUC | Weighted C-index | Weighted AUC | Weighted C-index |
| Clinicopathological | Breslow thickness*, ulceration | 0.82 (95% CI 0.78-0.86) | 0.85 (95% CI 0.72-0.88) | 0.82 (95% CI 0.73-0.90) | 0.84 (95% CI 0.76-0.91) |
| AJCC stage | Stage IA, IB, IIA, IIB, IIC | 0.79 (95% CI 0.75-0.82) | 0.83 (95% CI 0.79-0.86) | 0.80 (95% CI 0.70-0.89) | 0.81 (95% CI 0.72-0.89) |
| Matching variables | Age*, sex, Breslow thickness*, and ulceration | 0.79 (95% CI 0.74-0.83) | 0.83 (95% CI 0.78-0.86) | 0.81 (95% CI 0.72-0.89) | 0.83 (95% CI 0.75-0.90) |
| GEP | 558 discovery genes all tumors | 0.79 (95% CI 0.74-0.88) | 0.82 (95% CI 0.77-0.89) | 0.77 (95% CI 0.66-0.86) | 0.79 (95% CI 0.69-0.88) |
|  | 558 discovery genes in stage I | 0.72 (95% CI 0.54-0.81) | 0.72 (95% CI 0.54-0.82) | 0.69 (95% CI 0.52-0.84) | 0.64 (95% CI 0.46-0.80) |
|  | 558 discovery genes in stage II | 0.47 (95% CI 0.20-0.75) | 0.54 (95% CI 0.27-0.77) | 0.46 (95% CI 0.25-0.70) | 0.48 (95% CI 0.26-0.71) |
| Clinicopathological +<br>GEP | Breslow thickness*, ulceration,<br>558 genes from discovery | 0.80 (95% CI 0.74-0.88) | 0.83 (95% CI 0.77-0.89) | 0.79 (95% CI 0.68-0.87) | 0.81 (95% CI 0.71-0.89) |
| High selection<br>frequency GEP | 11 genes | 0.71 (95% CI 0.65-0.77) | 0.75 (95% CI 0.70-0.81) | 0.72 (95% CI 0.61-0.82) | 0.75 (95% CI 0.64-0.84) |

Various gene selection via regularization methods were explored, but neither adaptive LASSO nor ridge regression improved predictive accuracy. We also tested whether using subsets of genes with higher selection frequency in the discovery set (*n* = 11, 80, or 117) would enhance performance, but no improvement was observed (Table 2; Supplementary Figure 5).

### Independent validation

In the independent validation subset, we evaluated models developed in the model development subset. The clinical, gene-based, and integrated models showed similar performances; Breslow thickness and ulceration achieved a weighted AUC of 0.82 (95% CI, 0.73-0.90), and weighted C-index of 0.84 (95% CI, 0.76-0.91), the gene-only model showed a weighted AUC of 0.77 (95% CI, 0.66-0.86) and a weighted C-index of 0.79 (95% CI, 0.69-0.88); and the combined model showed a weighted AUC of 0.79 (95% CI, 0.68-0.87) and a weighted C-index of 0.81 (95% CI, 0.71-0.89) (Table 2, Figure 2B).

### Association of discovery genes with clinicopathological variables

To further explore the extent to which the 558 discovery genes reflected established prognostic variables, we first examined their direct association with Breslow thickness and ulceration. We observed that 75% of the genes were associated with ulceration and 79% with Breslow thickness in the discovery set. In the model development subset of the validation cohort, 96% of genes correlated with both variables. The aggregated prediction score based on the 558 genes showed a strong correlation with Breslow thickness (r = 0.74) and significantly higher values in ulcerated compared with non-ulcerated tumors (p = 1.1 × 10⁻¹⁴) (Supplementary Figure 6).

We then conducted principal component analysis (PCA) on the 558 discovery genes to further evaluate their independent contribution. PC1 was strongly associated with clinical stage, indicating overlap with known prognostic features (Supplementary Figure 7A). Adding clinical predictors to models based on the remaining PCs (excluding PC1) did not improve predictive performance, suggesting that GEP adds limited independent signal (Table 2, Figure 2). PCA-derived risk scores based on the remaining components appeared largely orthogonal to the clinical model, implying that gene expression may reflect distinct underlying biology (Supplementary Figure 7B).

### GEP models trained within the model development subset

GEP models trained within the model development subset (without relying on the 558 genes identified in the discovery set) showed comparable predictive performance in the independent validation subset. The best-performing model, based on 50 input features, achieved a weighted c-index of 0.81 (95% CI, 0.71-0.89) (Table 2). The results were comparable to those of the clinical model based on Breslow thickness and ulceration.

### Gene set enrichment analyses

Gene set enrichment analyses were performed in the model development to explore biological processes associated with metastatic progression. When conditioned on the matching variables, several hallmark gene sets were significantly enriched in metastatic cases. The top enriched pathways included E2F targets, MYC targets (V1), and the G2M checkpoint, all showing strong positive enrichment with high normalized enrichment scores (NES). Additional enrichment was seen for hypoxia, estrogen response, unfolded protein response, and both interferon gamma and alpha signaling (Figure 3A).

**Fig 3.**
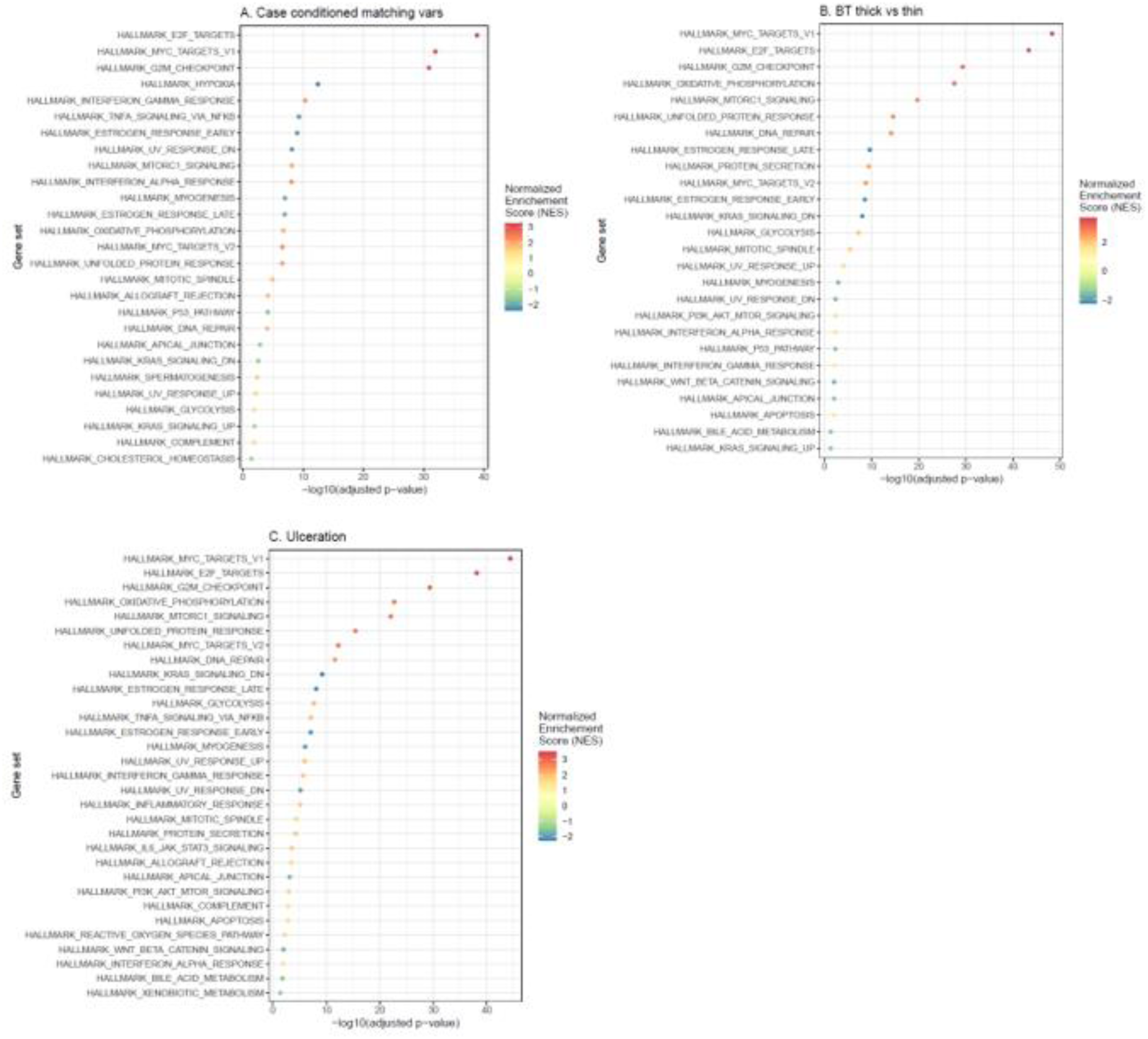
Gene set enrichment analyses (GSEA) of hallmark gene set in the model development subset of the validation cohort. A. Case-control analysis conditioned on matching variables (age, sex, Breslow thickness, ulceration). B. GSEA following differential expression analysis by Breslow thickness (>2 mm vs ≤2 mm). C. GSEA following differential expression analysis by ulceration status (present vs absent).

To assess whether these signals overlap with established clinicopathological risk factors, we repeated the analyses stratified by Breslow thickness and ulceration. In thicker melanomas (>2 mm), the same proliferative programs (MYC targets, E2F targets, G2M checkpoint) were again among the top enriched gene sets, accompanied by oxidative phosphorylation, DNA repair, and mTORC1 signaling (Figure 3B). Similarly, in ulcerated melanomas, a broad set of hallmark pathways was enriched, including proliferative signaling (E2F, MYC, G2M), metabolic activity (glycolysis, oxidative phosphorylation, fatty acid metabolism), and immune responses (inflammatory response, interferon signaling) (Figure 3C).

### Exploratory analyses of publicly available gene sets

We conducted exploratory analyses of previously published gene sets within the model development subset of the validation cohort ^7–11^. None of these sets showed a distinct differential expression pattern in volcano plots (Supplementary Figure 8). Predictive models built from these published gene sets demonstrated discriminative performance similar to that of our 558-gene set, and comparable to models based on Breslow thickness with ulceration or on AJCC stage (Figure 4).

**Fig 4.**
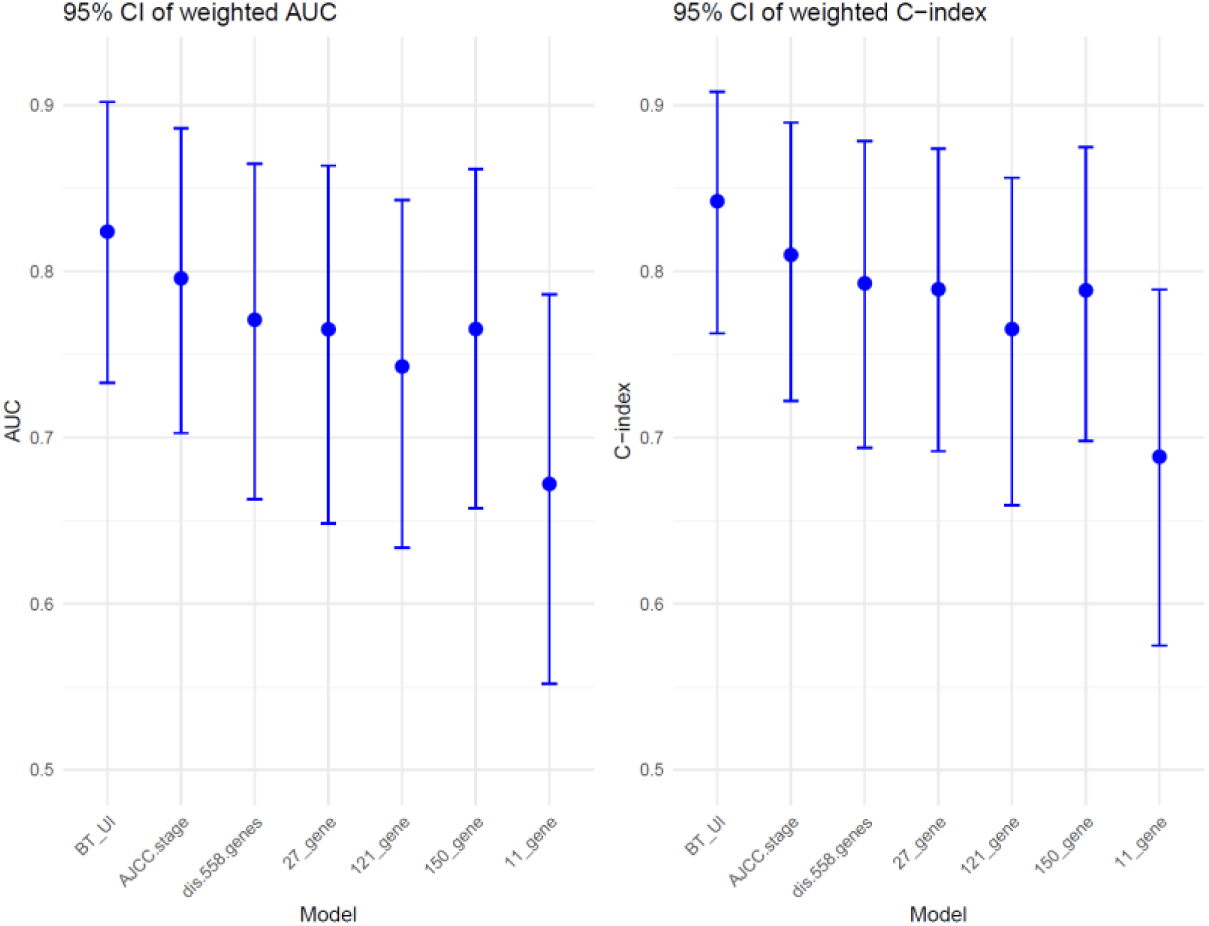
Weighted area under the curve (AUC (left) and weighted C-index (right) with 95% confidence intervals based on 1,000 bootstrap iterations (B = 1000) are shown for models based on Breslow thickness and ulceration (BT_Ul), AJCC stage, the 558 discovery genes, and previously published gene sets.

## Discussion

Accurately identifying early-stage melanoma patients at risk of distant metastases remains clinically challenging. Using data from the D-ESMEL study, we developed and validated GEP models for predicting distant metastases in stage I/II melanoma. While the GEP revealed biologically relevant transcriptional differences, its predictive performance for distant metastases was comparable to that of Breslow thickness and ulceration. These findings suggest that the prognostic information captured by gene expression largely overlaps with, and may partly reflect, established clinicopathological variables.

GEP assays have demonstrated significant promise in melanoma risk stratification. A recent systematic review and meta-analyses of the DecisionDx-Melanoma (Castle Biosciences Inc.) with >14,000 patients have demonstrated consistent risk stratification with significantly different survival outcomes ^30^. CP-GEP (SkylineDx) has been validated in multiple independent cohorts, with a recent systematic review and meta-analysis involving >1,000 patients demonstrating that CP-GEP performs well in identifying melanoma patients at low risk of SLN positivity, showing high negative predictive value (95%), supporting its value in SLNB decision-making ^31^. With ongoing prospective trials like NivoMela (NCT04309409), the utility of Melagenix (NeraCare GmbH) in guiding adjuvant therapy decisions is being evaluated. At the same time, translating GEP promise into consistent clinical utility faces methodological challenges, particularly in studies with relatively few outcome events. For example, population-based SEER data reported high hazard ratios for DecisionDx-Melanoma classifications, but with only 68/4,687 melanoma-specific deaths, the 95% CI’s were wide (HR 28.25 [95% CI: 12.69–62.89] univariable; HR 7.00 [95% CI: 2.70–18.00] multivariable) ^19^. Similarly, the LMC_150 signature was associated with melanoma-specific survival in stage I patients, but effect estimates were less precise after adjustment for clinicopathological variables (e.g., HR 9.8 [95% CI: 1.1–86.2]) ^9^. The Cam_121 signature, developed in stage II disease, showed minimal discrimination in the earlier-stage Leeds Melanoma Cohort, highlighting potential limitations in generalizability across stages (five-year melanoma-specific mortality 9% vs. 8%) ^10^. These examples highlight the importance of study design, outcome definitions, and patient selection when evaluating the added value of GEPs.

Our study design differs from prior GEP investigations by explicitly focusing on distant metastases as the primary endpoint, excluding patients with only local or regional relapse. This approach emphasizes the most clinically relevant outcome and leverages our population-based cohort with extended follow-up. As outlined in methodological guidance, prognostic models should be developed using adequately powered datasets with sufficient events per variable, include internal validation to quantify optimism, undergo external validation to assess generalizability, and demonstrate incremental value beyond established prognostic factors ^18,32^. The D-ESMEL study was designed to meet these criteria, including a matched discovery design to isolate signals beyond age, sex, Breslow thickness, and ulceration; and a large, population-based validation cohort. As an internal validation of our methodology, we further evaluated previously published melanoma gene sets. Although technical and clinical differences preclude direct performance comparisons, the concordant results we observed underscore the robustness of our dataset and analyses.

Several factors may explain why the discovery genes did not translate into stronger predictive models. Despite tight matching on Breslow thickness and ulceration in the discovery set, many genes remained strongly correlated with these factors. Matching balances variables at the cohort level but does not remove their intrinsic biological links with gene expression. In contrast, the validation cohort reflected a natural clinical distribution with more thin melanomas among controls. However, stratified analyses by AJCC stage showed comparable model performance across stage subgroups. Differences in SLNB uptake may also have played a role. Only 32% of discovery cases underwent SLNB, compared with 47% in the validation cohort, reflecting temporal shifts in clinical practice after the introduction of adjuvant therapy in 2019. Lower SLNB uptake may have resulted in under detection of nodal metastases. In contrast, higher SLNB rates in the validation cohort, likely improved staging accuracy, reducing the proportion of occult stage III cases. These differences in case composition, combined with the strong correlation between gene expression and clinicopathological features, likely account for their comparable predictive performance. Despite this overlap, our GEP model captured biologically relevant information. In the independent validation subset, the GEP models demonstrated solid performance (weighted C-index = 0.79).

Gene set enrichment analyses revealed consistent enrichment of proliferative programs, as well as hypoxia, estrogen response, unfolded protein response, and interferon signaling: pathways known to drive tumor aggressiveness ^33–37^. Importantly, the same pathways were enriched in thicker and ulcerated tumors, reinforcing the notion that the biology captured by GEP mirrors established high-risk clinicopathological features. In line with prior data, thicker tumors showed upregulation of DNA repair and cell-cycle genes ^38^, while ulcerated primaries exhibited proliferative, vascular, and inflammatory profiles ^39^. These findings reaffirm that the prognostic signal captured by GEP substantially overlaps with the biology already reflected by Breslow thickness and ulceration. Nonetheless, the biological insights remain valuable. GEP may support biological stratification in research settings, integration into multi-modal models combining molecular, spatial, or immune features, or provide supportive context in histologically ambiguous cases.

In summary, the D-ESMEL study provides a population-based, methodologically rigorous evaluation of GEP for predicting distant metastases in early-stage melanoma. Our findings reaffirm the dominant role of Breslow thickness and ulceration, show that GEP captures overlapping biology, and highlight key considerations for translating molecular data into clinically useful risk stratification.

## Data Availability

The clinical data used in this study were obtained from the Netherlands Cancer Registry (NCR) and are available through the Netherlands Comprehensive Cancer Organization. These data are not publicly available, and restrictions apply to their use. Access may be granted upon reasonable request and with permission from the Netherlands Comprehensive Cancer Organization (request numbers K19.037 and K21.277). The list of 558 candidate genes is provided in Supplementary Table 1. Gene expression data (counts per gene), including sample ID, pair ID, and case-control status will be made available via a public repository (pending).

## Acknowledgments

We thank all dedicated registrars of the NCR and Palga for their work on the data collection. We thank the Erasmus MC Pathology Research and Trial Service (PARTS) for sectioning the tissue material. The project was co-funded by the PPP Allowance made available by Health∼Holland (Grant Number: EMCLSH19008), Top Sector Life Sciences & Health.

## Data availability

The clinical data used in this study were obtained from the Netherlands Cancer Registry (NCR) and are available through the Netherlands Comprehensive Cancer Organization. These data are not publicly available, and restrictions apply to their use. Access may be granted upon reasonable request and with permission from the Netherlands Comprehensive Cancer Organization (request numbers K19.037 and K21.277). The list of 558 candidate genes is provided in Supplementary Table 1. Gene expression data (counts per gene), including sample ID, pair ID, and case-control status, will be made available via a public repository (pending).

## References

1. Gershenwald JE, Scolyer RA, Hess KR, et al: AJCC cancer staging manual. Switzerland: Springer:563–89, 2017

2. Zhou C, Louwman M, Wakkee M, et al: Primary Melanoma Characteristics of Metastatic Disease: A Nationwide Cancer Registry Study. Cancers (Basel) 13, 2021

3. Leeneman B, Franken MG, Coupé VMH, et al: Stage-specific disease recurrence and survival in localized and regionally advanced cutaneous melanoma. Eur. J. Surg. Oncol. 45:825–831, 2019

4. Enninga EAL, Moser JC, Weaver AL, et al: Survival of cutaneous melanoma based on sex, age, and stage in the United States, 1992-2011. Cancer Med. 6:2203–2212, 2017

5. Grossman D, Okwundu N, Bartlett EK, et al: Prognostic Gene Expression Profiling in Cutaneous Melanoma: Identifying the Knowledge Gaps and Assessing the Clinical Benefit. JAMA Dermatol 156:1004–1011, 2020

6. Sun J, Karasaki KM, Farma JM: The Use of Gene Expression Profiling and Biomarkers in Melanoma Diagnosis and Predicting Recurrence: Implications for Surveillance and Treatment. Cancers (Basel) 16, 2024

7. Meves A, Nikolova E, Heim JB, et al: Tumor Cell Adhesion As a Risk Factor for Sentinel Lymph Node Metastasis in Primary Cutaneous Melanoma. J. Clin. Oncol. 33:2509–15, 2015

8. Gerami P, Cook RW, Wilkinson J, et al: Development of a prognostic genetic signature to predict the metastatic risk associated with cutaneous melanoma. Clin. Cancer. Res. 21:175–83, 2015

9. Thakur R, Laye JP, Lauss M, et al: Transcriptomic Analysis Reveals Prognostic Molecular Signatures of Stage I Melanoma. Clin. Cancer Res. 25:7424–7435, 2019

10. Garg M, Couturier DL, Nsengimana J, et al: Tumour gene expression signature in primary melanoma predicts long-term outcomes. Nat. Commun. 12:1137, 2021

11. Brunner G, Heinecke A, Falk TM, et al: A Prognostic Gene Signature Expressed in Primary Cutaneous Melanoma: Synergism With Conventional Staging. JNCI Cancer Spectr 2:pky032, 2018

12. Bellomo D, Arias-Mejias SM, Ramana C, et al: Model Combining Tumor Molecular and Clinicopathologic Risk Factors Predicts Sentinel Lymph Node Metastasis in Primary Cutaneous Melanoma. JCO Precis. Oncol. 4:319–334, 2020

13. Mulder E, Dwarkasing JT, Tempel D, et al: Validation of a clinicopathological and gene expression profile model for sentinel lymph node metastasis in primary cutaneous melanoma. Br J Dermatol 184:944–951, 2021

14. Stassen RC, Mulder E, Mooyaart AL, et al: Clinical evaluation of the clinicopathologic and gene expression profile (CP-GEP) in patients with melanoma eligible for sentinel lymph node biopsy: A multicenter prospective Dutch study. Eur J Surg Oncol 49:107249, 2023

15. National Comprehensive Cancer Network Clinical Practice Guidelines in Oncology Melanoma: Version 2.2025. , 2025

16. Bartlett EK, O’Donoghue C, Boland G, et al: Society of Surgical Oncology Consensus Statement: Assessing the Evidence for and Utility of Gene Expression Profiling of Primary Cutaneous Melanoma. Ann Surg Oncol 32:1429–1442, 2025

17. Swetter SM, Tsao H, Bichakjian CK, et al: Guidelines of care for the management of primary cutaneous melanoma. J. Am. Acad. Dermatol. 80:208–250, 2019

18. Steyerberg E: Clinical Prediction Models: A Practical Approach to Development, Validation, and Updating, Springer, 2019

19. Bailey CN, Martin BJ, Petkov VI, et al: 31-Gene Expression Profile Testing in Cutaneous Melanoma and Survival Outcomes in a Population-Based Analysis: A SEER Collaboration. JCO Precis Oncol 7:e2300044, 2023

20. Zhou C, Mooyaart AL, Kerkour T, et al: The Dutch Early-Stage Melanoma (D-ESMEL) study: a discovery set and validation cohort to predict the absolute risk of distant metastases in stage I/II cutaneous melanoma. Eur J Epidemiol 40:27–42, 2025

21. Rentroia-Pacheco B, Bellomo D, Lakeman IMM, et al: Weighted metrics are required when evaluating the performance of prediction models in nested case-control studies. BMC Med. Res. Methodol. 24:115, 2024

22. Noordzij M, van Diepen M, Caskey FC, Jager KJ: Relative risk versus absolute risk: one cannot be interpreted without the other. Nephrol. Dial. Transplant 32:ii13–ii18, 2017

23. Rentroia-Pacheco B, Steijlen O, Bellomo D, et al: Nested Case-Control and Case-cohort: efficient study designs to develop biomarker-based prediction models for rare outcomes. J Invest Dermatol, Submitted

24. Netherlands Code of Conduct for Research Integrity, The Association of Universities in the Netherlands, 2018

25. Tibshirani R: Regression Shrinkage and Selection via the Lasso. Journal of the Royal Statistical Society Series B (Methodological) 58:267–288, 1996

26. Love MI, Huber W, Anders S: Moderated estimation of fold change and dispersion for RNA-seq data with DESeq2. Genome Biol 15:550, 2014

27. Ritchie ME, Phipson B, Wu D, et al: limma powers differential expression analyses for RNA-sequencing and microarray studies. Nucleic Acids Res 43:e47, 2015

28. Efron B, Tibshirani R: Improvements on Cross-Validation: The .632+ Bootstrap Method.

29. Liberzon A, Birger C, Thorvaldsdóttir H, et al: The Molecular Signatures Database (MSigDB) hallmark gene set collection. Cell Syst 1:417–425, 2015

30. Durgham RA, Nassar SI, Gun R, et al: The Prognostic Value of the 31-Gene Expression Profile Test in Cutaneous Melanoma: A Systematic Review and Meta-Analysis. Cancers (Basel) 16, 2024

31. Wong T, Ch’ng S, Ferguson P, et al: Predictive performance of the clinicopathologic gene expression profile (CP-GEP) in identifying cutaneous melanoma patients for whom sentinel lymph node biopsy is unnecessary: A systematic review and meta-analysis. Crit Rev Oncol Hematol 214:104816, 2025

32. Collins GS, Reitsma JB, Altman DG, Moons KG: Transparent reporting of a multivariable prediction model for individual prognosis or diagnosis (TRIPOD): the TRIPOD statement. Bmj 350:g7594, 2015

33. Alla V, Engelmann D, Niemetz A, et al: E2F1 in melanoma progression and metastasis. J Natl Cancer Inst 102:127–33, 2010

34. Ross DA, Wilson GD: Expression of c-myc oncoprotein represents a new prognostic marker in cutaneous melanoma. Br J Surg 85:46–51, 1998

35. Cubillos-Ruiz JR, Bettigole SE, Glimcher LH: Tumorigenic and Immunosuppressive Effects of Endoplasmic Reticulum Stress in Cancer. Cell 168:692–706, 2017

36. Di Conza G, Ho PC, Cubillos-Ruiz JR, Huang SC: Control of immune cell function by the unfolded protein response. Nat Rev Immunol 23:546–562, 2023

37. Grasso CS, Tsoi J, Onyshchenko M, et al: Conserved Interferon-γ Signaling Drives Clinical Response to Immune Checkpoint Blockade Therapy in Melanoma. Cancer Cell 38:500–515 e3, 2020

38. Fang S, Vaysse A, Brossard M, et al: Melanoma Expression Genes Identified through Genome-Wide Association Study of Breslow Tumor Thickness. J Invest Dermatol 137:253–257, 2017

39. Jewell R, Elliott F, Laye J, et al: The clinicopathological and gene expression patterns associated with ulceration of primary melanoma. Pigment Cell Melanoma Res 28:94–104, 2015

