## Supplementary Table 1 for "Gene expression profiling beyond Breslow thickness and ulceration for prediction of distant metastases in early-stage melanoma: the population-based Dutch Early-Stage Melanoma (D-ESMEL) study"

Supplementary Table 1: List of 558 genes. Genes were selected based on differential expression analysis and conditional logistic regression in the discovery set. Abbreviations: DLCV = double-loop cross-validation, DGEA = differential gene expression analyses

| Gene symbol | Ensembl ID | Source of selection |
|---|---|---|
| ANLN | ENSG00000011426.11 | DLCV and DGEA |
| OPN3 | ENSG00000054277.14 | DLCV and DGEA |
| NGFR | ENSG00000064300.9 | DLCV and DGEA |
| ATP12A | ENSG00000075673.12 | DLCV and DGEA |
| CFAP61 | ENSG00000089101.19 | DLCV and DGEA |
| OCA2 | ENSG00000104044.16 | DLCV and DGEA |
| EYA1 | ENSG00000104313.20 | DLCV and DGEA |
| CALB1 | ENSG00000104327.7 | DLCV and DGEA |
| CENPF | ENSG00000117724.13 | DLCV and DGEA |
| PPP1R3C | ENSG00000119938.9 | DLCV and DGEA |
| TSFM | ENSG00000123297.20 | DLCV and DGEA |
| HJURP | ENSG00000123485.12 | DLCV and DGEA |
| IL13RA2 | ENSG00000123496.8 | DLCV and DGEA |
| H1-1 | ENSG00000124610.5 | DLCV and DGEA |
| FGF13 | ENSG00000129682.16 | DLCV and DGEA |
| TNNI2 | ENSG00000130598.16 | DLCV and DGEA |
| MNX1 | ENSG00000130675.15 | DLCV and DGEA |
| KRT33B | ENSG00000131738.11 | DLCV and DGEA |
| GPR12 | ENSG00000132975.8 | DLCV and DGEA |
| SERPINE2 | ENSG00000135919.14 | DLCV and DGEA |
| EFCAB11 | ENSG00000140025.16 | DLCV and DGEA |
| ADAD2 | ENSG00000140955.11 | DLCV and DGEA |
| CELA2A | ENSG00000142615.8 | DLCV and DGEA |
| NBPF3 | ENSG00000142794.19 | DLCV and DGEA |
| COL8A1 | ENSG00000144810.16 | DLCV and DGEA |
| ANO4 | ENSG00000151572.18 | DLCV and DGEA |
| PSTPIP2 | ENSG00000152229.18 | DLCV and DGEA |
| ARSL | ENSG00000157399.17 | DLCV and DGEA |
| NRG2 | ENSG00000158458.21 | DLCV and DGEA |
| PM20D1 | ENSG00000162877.13 | DLCV and DGEA |
| RFTN2 | ENSG00000162944.11 | DLCV and DGEA |
| SPATA18 | ENSG00000163071.11 | DLCV and DGEA |
| GRM2 | ENSG00000164082.15 | DLCV and DGEA |
| NPY1R | ENSG00000164128.7 | DLCV and DGEA |
| CHMP4C | ENSG00000164695.5 | DLCV and DGEA |
| CMTM5 | ENSG00000166091.21 | DLCV and DGEA |
| WDR72 | ENSG00000166415.15 | DLCV and DGEA |
| TMEM170A | ENSG00000166822.13 | DLCV and DGEA |
| GTSF1 | ENSG00000170627.11 | DLCV and DGEA |
| HS6ST2 | ENSG00000171004.18 | DLCV and DGEA |
| ANGPTL7 | ENSG00000171819.5 | DLCV and DGEA |
| FRMD5 | ENSG00000171877.21 | DLCV and DGEA |

| | | |
|---|---|---|
| TPSAB1 | ENSG00000172236.18 | DLCV and DGEA |
| COL6A5 | ENSG00000172752.14 | DLCV and DGEA |
| KRT2 | ENSG00000172867.4 | DLCV and DGEA |
| C11orf45 | ENSG00000174370.11 | DLCV and DGEA |
| GCSAM | ENSG00000174500.13 | DLCV and DGEA |
| HOXD8 | ENSG00000175879.9 | DLCV and DGEA |
| GLDC | ENSG00000178445.10 | DLCV and DGEA |
| WASHC1 | ENSG00000181404.17 | DLCV and DGEA |
| PHLDA2 | ENSG00000181649.8 | DLCV and DGEA |
| KRTAP11-1 | ENSG00000182591.6 | DLCV and DGEA |
| CADM1 | ENSG00000182985.18 | DLCV and DGEA |
| CTAG1B | ENSG00000184033.14 | DLCV and DGEA |
| S100A3 | ENSG00000188015.10 | DLCV and DGEA |
| SERPINB2 | ENSG00000197632.9 | DLCV and DGEA |
| RYR2 | ENSG00000198626.17 | DLCV and DGEA |
| SHISA4 | ENSG00000198892.7 | DLCV and DGEA |
| RNU4-1 | ENSG00000200795.1 | DLCV and DGEA |
| HSD3B1 | ENSG00000203857.10 | DLCV and DGEA |
| MT1H | ENSG00000205358.4 | DLCV and DGEA |
| KRTAP10-2 | ENSG00000205445.3 | DLCV and DGEA |
| KRTAP2-1 | ENSG00000212725.3 | DLCV and DGEA |
| KRTAP4-12 | ENSG00000213416.4 | DLCV and DGEA |
| KRTAP2-4 | ENSG00000213417.3 | DLCV and DGEA |
| TTLL13P | ENSG00000213471.10 | DLCV and DGEA |
| KRTAP1-5 | ENSG00000221852.5 | DLCV and DGEA |
| AC079807.2 | ENSG00000233230.1 | DLCV and DGEA |
| WWTR1-AS1 | ENSG00000241313.3 | DLCV and DGEA |
| RP11-688I9.2 | ENSG00000255317.1 | DLCV and DGEA |
| BCDIN3D-AS1 | ENSG00000258057.6 | DLCV and DGEA |
| DUX4 | ENSG00000260596.5 | DLCV and DGEA |
| OPN1MW | ENSG00000268221.6 | DLCV and DGEA |
| SYNPO2L-AS1 | ENSG00000271848.2 | DLCV and DGEA |
| RP11-96C23.5 | ENSG00000271880.1 | DLCV and DGEA |
| RP11-834C11.12 | ENSG00000273049.1 | DLCV and DGEA |
| RP11-437B10.1 | ENSG00000279765.3 | DLCV and DGEA |
| SLURP2 | ENSG00000283992.2 | DLCV and DGEA |
| RP5-1091N2.12 | ENSG00000285171.1 | DLCV and DGEA |
| RNY5 | ENSG00000286171.1 | DLCV and DGEA |
| TNMD | ENSG00000000005.6 | DLCV |
| KRT33A | ENSG00000006059.4 | DLCV |
| TNFRSF12A | ENSG00000006327.14 | DLCV |
| SYN1 | ENSG00000008056.14 | DLCV |
| ISL1 | ENSG00000016082.15 | DLCV |
| IGF1 | ENSG00000017427.17 | DLCV |
| DEPDC1 | ENSG00000024526.17 | DLCV |
| DEPDC1B | ENSG00000035499.13 | DLCV |

| | | |
|---|---|---|
| CDH10 | ENSG00000040731.10 | DLCV |
| ANO2 | ENSG00000047617.16 | DLCV |
| TNFRSF9 | ENSG00000049249.9 | DLCV |
| CCN5 | ENSG00000064205.11 | DLCV |
| SNX24 | ENSG00000064652.11 | DLCV |
| CAMK2A | ENSG00000070808.17 | DLCV |
| SNCB | ENSG00000074317.11 | DLCV |
| NUAK1 | ENSG00000074590.14 | DLCV |
| SCTR | ENSG00000080293.10 | DLCV |
| CHRNA3 | ENSG00000080644.16 | DLCV |
| MRPL22 | ENSG00000082515.18 | DLCV |
| HAL | ENSG00000084110.11 | DLCV |
| COL16A1 | ENSG00000084636.18 | DLCV |
| SMPX | ENSG00000091482.8 | DLCV |
| CDC45 | ENSG00000093009.11 | DLCV |
| GABRP | ENSG00000094755.17 | DLCV |
| CRISP3 | ENSG00000096006.12 | DLCV |
| CDC7 | ENSG00000097046.13 | DLCV |
| GGT1 | ENSG00000100031.19 | DLCV |
| RSPH14 | ENSG00000100218.12 | DLCV |
| CGRRF1 | ENSG00000100532.13 | DLCV |
| LRRC74A | ENSG00000100565.15 | DLCV |
| BDKRB1 | ENSG00000100739.11 | DLCV |
| SLC52A3 | ENSG00000101276.18 | DLCV |
| STMN2 | ENSG00000104435.14 | DLCV |
| CASP14 | ENSG00000105141.6 | DLCV |
| ICAM5 | ENSG00000105376.5 | DLCV |
| CCL24 | ENSG00000106178.7 | DLCV |
| SERPINE1 | ENSG00000106366.9 | DLCV |
| PSMA2 | ENSG00000106588.12 | DLCV |
| DNAJC12 | ENSG00000108176.15 | DLCV |
| MPP2 | ENSG00000108852.15 | DLCV |
| SLC16A6 | ENSG00000108932.12 | DLCV |
| SULT1E1 | ENSG00000109193.12 | DLCV |
| CDCA3 | ENSG00000111665.12 | DLCV |
| SIM1 | ENSG00000112246.10 | DLCV |
| UNC93A | ENSG00000112494.10 | DLCV |
| HBEGF | ENSG00000113070.8 | DLCV |
| PCDHB5 | ENSG00000113209.9 | DLCV |
| FGF1 | ENSG00000113578.18 | DLCV |
| IL1R2 | ENSG00000115590.14 | DLCV |
| PLEK | ENSG00000115956.10 | DLCV |
| OLFML3 | ENSG00000116774.12 | DLCV |
| MFAP2 | ENSG00000117122.14 | DLCV |
| ARG1 | ENSG00000118520.15 | DLCV |
| GDA | ENSG00000119125.17 | DLCV |

| | | |
|---|---|---|
| GRIA2 | ENSG00000120251.21 | DLCV |
| TP53AIP1 | ENSG00000120471.16 | DLCV |
| PLS1 | ENSG00000120756.13 | DLCV |
| ADRA1A | ENSG00000120907.18 | DLCV |
| CSTA | ENSG00000121552.4 | DLCV |
| CCR2 | ENSG00000121807.7 | DLCV |
| HS3ST2 | ENSG00000122254.7 | DLCV |
| CIT | ENSG00000122966.17 | DLCV |
| ACVR1C | ENSG00000123612.16 | DLCV |
| IL9R | ENSG00000124334.17 | DLCV |
| HCST | ENSG00000126264.10 | DLCV |
| CTAG2 | ENSG00000126890.14 | DLCV |
| APOBEC3A | ENSG00000128383.13 | DLCV |
| OPN1SW | ENSG00000128617.3 | DLCV |
| CDKN1C | ENSG00000129757.15 | DLCV |
| ATP8B3 | ENSG00000130270.16 | DLCV |
| ACSBG2 | ENSG00000130377.14 | DLCV |
| RAMP1 | ENSG00000132329.11 | DLCV |
| CPLANE2 | ENSG00000132881.12 | DLCV |
| DCLK1 | ENSG00000133083.15 | DLCV |
| RSAD2 | ENSG00000134321.13 | DLCV |
| ADAMTS8 | ENSG00000134917.10 | DLCV |
| DRAM1 | ENSG00000136048.14 | DLCV |
| CIDEB | ENSG00000136305.11 | DLCV |
| CHAD | ENSG00000136457.10 | DLCV |
| FRS3 | ENSG00000137218.10 | DLCV |
| FXYD6 | ENSG00000137726.17 | DLCV |
| AOX1 | ENSG00000138356.14 | DLCV |
| FBN2 | ENSG00000138829.12 | DLCV |
| LGR5 | ENSG00000139292.13 | DLCV |
| MORN3 | ENSG00000139714.12 | DLCV |
| CBLN3 | ENSG00000139899.11 | DLCV |
| MYOCD | ENSG00000141052.18 | DLCV |
| SKAP1 | ENSG00000141293.16 | DLCV |
| P3H4 | ENSG00000141696.13 | DLCV |
| FKBP10 | ENSG00000141756.19 | DLCV |
| FLG | ENSG00000143631.11 | DLCV |
| DLX1 | ENSG00000144355.15 | DLCV |
| TRPM8 | ENSG00000144481.17 | DLCV |
| OSBPL10 | ENSG00000144645.15 | DLCV |
| MED12L | ENSG00000144893.12 | DLCV |
| MUC4 | ENSG00000145113.22 | DLCV |
| CORIN | ENSG00000145244.12 | DLCV |
| CPLX2 | ENSG00000145920.15 | DLCV |
| TIAM2 | ENSG00000146426.19 | DLCV |
| CDCA5 | ENSG00000146670.10 | DLCV |

| | | |
|---|---|---|
| TRIM50 | ENSG00000146755.11 | DLCV |
| GAS2 | ENSG00000148935.11 | DLCV |
| HMGA2 | ENSG00000149948.14 | DLCV |
| FREM2 | ENSG00000150893.11 | DLCV |
| AP1S3 | ENSG00000152056.17 | DLCV |
| ANKRD22 | ENSG00000152766.6 | DLCV |
| PTPRD | ENSG00000153707.18 | DLCV |
| SDHAF4 | ENSG00000154079.6 | DLCV |
| EPHB1 | ENSG00000154928.19 | DLCV |
| ADCY8 | ENSG00000155897.10 | DLCV |
| ALX3 | ENSG00000156150.9 | DLCV |
| CD1C | ENSG00000158481.13 | DLCV |
| SIM2 | ENSG00000159263.16 | DLCV |
| HSF2BP | ENSG00000160207.9 | DLCV |
| TMEM143 | ENSG00000161558.11 | DLCV |
| CD300LG | ENSG00000161649.13 | DLCV |
| SPC24 | ENSG00000161888.11 | DLCV |
| SPATA17 | ENSG00000162814.11 | DLCV |
| CCDC74A | ENSG00000163040.15 | DLCV |
| BBS5 | ENSG00000163093.12 | DLCV |
| NIPAL1 | ENSG00000163293.12 | DLCV |
| SERPINI1 | ENSG00000163536.13 | DLCV |
| FANCD2OS | ENSG00000163705.13 | DLCV |
| IL31RA | ENSG00000164509.16 | DLCV |
| SMCO2 | ENSG00000165935.9 | DLCV |
| C2 | ENSG00000166278.15 | DLCV |
| BEAN1 | ENSG00000166546.14 | DLCV |
| PROCA1 | ENSG00000167525.15 | DLCV |
| C19orf33 | ENSG00000167644.12 | DLCV |
| PLAAT5 | ENSG00000168004.10 | DLCV |
| WFDC12 | ENSG00000168703.5 | DLCV |
| LINC01620 | ENSG00000168746.8 | DLCV |
| VXN | ENSG00000169085.13 | DLCV |
| GSG1L | ENSG00000169181.13 | DLCV |
| GON7 | ENSG00000170270.5 | DLCV |
| KRT4 | ENSG00000170477.13 | DLCV |
| RAB37 | ENSG00000172794.20 | DLCV |
| CCR9 | ENSG00000173585.17 | DLCV |
| RGMB | ENSG00000174136.13 | DLCV |
| TNK1 | ENSG00000174292.12 | DLCV |
| FOSL1 | ENSG00000175592.9 | DLCV |
| DOK7 | ENSG00000175920.18 | DLCV |
| MUC20 | ENSG00000176945.17 | DLCV |
| ZFAS1 | ENSG00000177410.13 | DLCV |
| MBOAT4 | ENSG00000177669.4 | DLCV |
| DNAJC22 | ENSG00000178401.16 | DLCV |

| | | |
|---|---|---|
| C9orf50 | ENSG00000179058.7 | DLCV |
| C12orf42 | ENSG00000179088.15 | DLCV |
| VWA1 | ENSG00000179403.12 | DLCV |
| LACC1 | ENSG00000179630.11 | DLCV |
| GOLGA8J | ENSG00000179938.12 | DLCV |
| ITPRID1 | ENSG00000180347.14 | DLCV |
| OR52N4 | ENSG00000181074.5 | DLCV |
| OTOP2 | ENSG00000183034.13 | DLCV |
| TREX2 | ENSG00000183479.13 | DLCV |
| SFXN4 | ENSG00000183605.17 | DLCV |
| CFAP91 | ENSG00000183833.16 | DLCV |
| FAM3B | ENSG00000183844.17 | DLCV |
| SRPK3 | ENSG00000184343.11 | DLCV |
| OSBP2 | ENSG00000184792.16 | DLCV |
| FAM227A | ENSG00000184949.18 | DLCV |
| ROBO2 | ENSG00000185008.17 | DLCV |
| MROH2A | ENSG00000185038.14 | DLCV |
| CDNF | ENSG00000185267.10 | DLCV |
| CYP4F12 | ENSG00000186204.15 | DLCV |
| MTARC1 | ENSG00000186205.13 | DLCV |
| CD300E | ENSG00000186407.7 | DLCV |
| CDHR4 | ENSG00000187492.9 | DLCV |
| CCDC157 | ENSG00000187860.11 | DLCV |
| FAM166A | ENSG00000188163.8 | DLCV |
| KRTDAP | ENSG00000188508.11 | DLCV |
| DUSP28 | ENSG00000188542.10 | DLCV |
| ALKAL2 | ENSG00000189292.16 | DLCV |
| ASMT | ENSG00000196433.13 | DLCV |
| CACNA1H | ENSG00000196557.13 | DLCV |
| ZNF239 | ENSG00000196793.14 | DLCV |
| GAL3ST4 | ENSG00000197093.11 | DLCV |
| C6orf141 | ENSG00000197261.11 | DLCV |
| STPG3 | ENSG00000197768.10 | DLCV |
| SLC34A3 | ENSG00000198569.10 | DLCV |
| RNU5A-1 | ENSG00000199568.1 | DLCV |
| MICB | ENSG00000204516.10 | DLCV |
| ASPDH | ENSG00000204653.10 | DLCV |
| ODAD4 | ENSG00000204815.10 | DLCV |
| SPDYE2 | ENSG00000205238.10 | DLCV |
| CDPF1 | ENSG00000205643.11 | DLCV |
| KRTAP5-1 | ENSG00000205869.2 | DLCV |
| HCG27 | ENSG00000206344.7 | DLCV |
| KLK9 | ENSG00000213022.6 | DLCV |
| MAGEA12 | ENSG00000213401.10 | DLCV |
| ZBTB9 | ENSG00000213588.6 | DLCV |
| SLC23A3 | ENSG00000213901.11 | DLCV |

| | | |
|---|---|---|
| CCL27 | ENSG00000213927.4 | DLCV |
| SMIM7 | ENSG00000214046.9 | DLCV |
| MYCBP | ENSG00000214114.9 | DLCV |
| TMEM213 | ENSG00000214128.11 | DLCV |
| C19orf38 | ENSG00000214212.9 | DLCV |
| SOGA3 | ENSG00000214338.10 | DLCV |
| AC087491.2 | ENSG00000214546.4 | DLCV |
| MTCP1 | ENSG00000214827.11 | DLCV |
| KRTAP10-1 | ENSG00000215455.4 | DLCV |
| KRTAP1-3 | ENSG00000221880.4 | DLCV |
| C19orf73 | ENSG00000221916.4 | DLCV |
| AC106873.4 | ENSG00000228368.1 | DLCV |
| ARL17B | ENSG00000228696.10 | DLCV |
| WEE2-AS1 | ENSG00000228775.8 | DLCV |
| THAP7-AS1 | ENSG00000230513.2 | DLCV |
| RP11-25K21.1 | ENSG00000234211.2 | DLCV |
| CNN3-DT | ENSG00000235501.6 | DLCV |
| MORC2-AS1 | ENSG00000235989.4 | DLCV |
| TAS2R39 | ENSG00000236398.2 | DLCV |
| GFOD1-AS1 | ENSG00000237786.1 | DLCV |
| OR2W3 | ENSG00000238243.3 | DLCV |
| TEX35 | ENSG00000240021.10 | DLCV |
| FOXO3B | ENSG00000240445.6 | DLCV |
| KRTAP4-7 | ENSG00000240871.5 | DLCV |
| RP11-111H13.1 | ENSG00000241962.9 | DLCV |
| PEG10 | ENSG00000242265.6 | DLCV |
| GLYCTK-AS1 | ENSG00000242797.3 | DLCV |
| SIAH2-AS1 | ENSG00000244265.2 | DLCV |
| ATP5MGL | ENSG00000249222.1 | DLCV |
| TRMT9B | ENSG00000250305.9 | DLCV |
| HOXA10 | ENSG00000253293.5 | DLCV |
| AC006277.2 | ENSG00000253392.3 | DLCV |
| CTD-2517M22.14 | ENSG00000255182.2 | DLCV |
| CYP2A6 | ENSG00000255974.8 | DLCV |
| C17orf100 | ENSG00000256806.6 | DLCV |
| RP11-284H19.1 | ENSG00000256879.1 | DLCV |
| RP11-454K7.1 | ENSG00000257900.2 | DLCV |
| RP11-649E7.5 | ENSG00000258377.1 | DLCV |
| RP11-84C10.4 | ENSG00000258471.2 | DLCV |
| CTD-2555O16.2 | ENSG00000258824.2 | DLCV |
| RP11-348B17.1 | ENSG00000259720.2 | DLCV |
| SLC22A31 | ENSG00000259803.8 | DLCV |
| ANKRD20A1 | ENSG00000260691.7 | DLCV |
| RP11-2C24.4 | ENSG00000260899.1 | DLCV |
| LMO7-AS1 | ENSG00000261105.6 | DLCV |
| TMEM178B | ENSG00000261115.6 | DLCV |

| | | |
|---|---|---|
| RP11-296A16.1 | ENSG00000262560.1 | DLCV |
| RP11-242D8.1 | ENSG00000267002.4 | DLCV |
| AC005262.3 | ENSG00000267139.1 | DLCV |
| CTB-54O9.9 | ENSG00000267157.1 | DLCV |
| RP11-318A15.7 | ENSG00000267168.1 | DLCV |
| RP11-1151B14.4 | ENSG00000267257.1 | DLCV |
| AC005306.3 | ENSG00000267283.1 | DLCV |
| SMIM22 | ENSG00000267795.6 | DLCV |
| CTD-2583A14.10 | ENSG00000268750.7 | DLCV |
| CSAG3 | ENSG00000268916.6 | DLCV |
| CTD-3099C6.5 | ENSG00000269349.1 | DLCV |
| CTD-2616J11.11 | ENSG00000269403.1 | DLCV |
| BIVM-ERCC5 | ENSG00000270181.3 | DLCV |
| TLCD4-RWDD3 | ENSG00000271092.5 | DLCV |
| MROH7-TTC4 | ENSG00000271723.5 | DLCV |
| KRTAP10-7 | ENSG00000272804.3 | DLCV |
| CTA-384D8.36 | ENSG00000272821.1 | DLCV |
| ENSG10010139146.1 | ENSG00000273658.1 | DLCV |
| RP11-353N4.6 | ENSG00000275557.1 | DLCV |
| RNU12 | ENSG00000276027.1 | DLCV |
| F8A3 | ENSG00000277150.2 | DLCV |
| ABC7-42404400C24.1 | ENSG00000277758.6 | DLCV |
| Telomerase-vert | ENSG00000277925.1 | DLCV |
| CH17-140K24.7 | ENSG00000278936.1 | DLCV |
| RP11-417N10.4 | ENSG00000279592.1 | DLCV |
| RP11-654K19.6 | ENSG00000285218.1 | DLCV |
| RP11-732A19.10 | ENSG00000285338.1 | DLCV |
| CTD-2308N23.4 | ENSG00000285347.1 | DLCV |
| RP11-468E2.12 | ENSG00000285467.1 | DLCV |
| RP11-79P21.2 | ENSG00000285708.1 | DLCV |
| F8A2 | ENSG00000288709.1 | DLCV |
| RP5-973N23.5 | ENSG00000288721.1 | DLCV |
| CDK6 | ENSG00000105810.10 | DGEA |
| TRIM24 | ENSG00000122779.18 | DGEA |
| PTPN12 | ENSG00000127947.16 | DGEA |
| SEC14L1 | ENSG00000129657.16 | DGEA |
| STX11 | ENSG00000135604.10 | DGEA |
| RAB15 | ENSG00000139998.16 | DGEA |
| SLC13A4 | ENSG00000164707.16 | DGEA |
| ALOXE3 | ENSG00000179148.10 | DGEA |
| PLEKHM1 | ENSG00000225190.11 | DGEA |
| FAM133B | ENSG00000234545.8 | DGEA |
| FOXM1 | ENSG00000111206.13 | DGEA |
| FNBP1 | ENSG00000187239.17 | DGEA |
| EPPK1 | ENSG00000261150.3 | DGEA |
| VPS9D1-AS1 | ENSG00000261373.1 | DGEA |

| | | |
|---|---|---|
| MYCBP2 | ENSG00000005810.19 | DGEA |
| EXOSC7 | ENSG00000075914.13 | DGEA |
| LAMB1 | ENSG00000091136.15 | DGEA |
| PTGS1 | ENSG00000095303.17 | DGEA |
| TRERF1 | ENSG00000124496.12 | DGEA |
| WBP2 | ENSG00000132471.12 | DGEA |
| SEC16A | ENSG00000148396.18 | DGEA |
| MPP7 | ENSG00000150054.19 | DGEA |
| CFDP1 | ENSG00000153774.9 | DGEA |
| TSC1 | ENSG00000165699.15 | DGEA |
| CNBP | ENSG00000169714.17 | DGEA |
| MAF | ENSG00000178573.7 | DGEA |
| GLRX5 | ENSG00000182512.5 | DGEA |
| KRT10 | ENSG00000186395.9 | DGEA |
| DBF4 | ENSG00000006634.8 | DGEA |
| SNX13 | ENSG00000071189.21 | DGEA |
| AURKA | ENSG00000087586.18 | DGEA |
| VIPR1 | ENSG00000114812.13 | DGEA |
| TOP2A | ENSG00000131747.15 | DGEA |
| ZFHX2 | ENSG00000136367.14 | DGEA |
| FADS1 | ENSG00000149485.19 | DGEA |
| ZCCHC10 | ENSG00000155329.12 | DGEA |
| BRD7 | ENSG00000166164.16 | DGEA |
| ANAPC2 | ENSG00000176248.9 | DGEA |
| SLC26A11 | ENSG00000181045.15 | DGEA |
| TNFAIP8L3 | ENSG00000183578.8 | DGEA |
| ZNF311 | ENSG00000197935.7 | DGEA |
| ELOVL2 | ENSG00000197977.4 | DGEA |
| VGLL3 | ENSG00000206538.9 | DGEA |
| EEF2K | ENSG00000103319.12 | DGEA |
| RAPGEF1 | ENSG00000107263.19 | DGEA |
| KNSTRN | ENSG00000128944.13 | DGEA |
| ERMN | ENSG00000136541.15 | DGEA |
| PIM1 | ENSG00000137193.14 | DGEA |
| ETV6 | ENSG00000139083.11 | DGEA |
| SCUBE3 | ENSG00000146197.9 | DGEA |
| CDHR1 | ENSG00000148600.15 | DGEA |
| SYT8 | ENSG00000149043.16 | DGEA |
| RAB11FIP1 | ENSG00000156675.16 | DGEA |
| VCP | ENSG00000165280.18 | DGEA |
| COG1 | ENSG00000166685.12 | DGEA |
| B4GALNT2 | ENSG00000167080.9 | DGEA |
| ZBTB43 | ENSG00000169155.10 | DGEA |
| HOPX | ENSG00000171476.22 | DGEA |
| CES4A | ENSG00000172824.16 | DGEA |
| RIMS2 | ENSG00000176406.23 | DGEA |

| | | |
|---|---|---|
| ADGRB1 | ENSG00000181790.11 | DGEA |
| NLRP10 | ENSG00000182261.4 | DGEA |
| PARPBP | ENSG00000185480.12 | DGEA |
| ANKRD35 | ENSG00000198483.13 | DGEA |
| RP1-20C7.6 | ENSG00000272223.1 | DGEA |
| PCGF2 | ENSG00000277258.5 | DGEA |
| TF | ENSG00000091513.16 | DGEA |
| IL11 | ENSG00000095752.7 | DGEA |
| SEC14L2 | ENSG00000100003.18 | DGEA |
| PLAT | ENSG00000104368.19 | DGEA |
| ATP4A | ENSG00000105675.9 | DGEA |
| TFPI2 | ENSG00000105825.14 | DGEA |
| EBF3 | ENSG00000108001.16 | DGEA |
| MYH1 | ENSG00000109061.10 | DGEA |
| MAK | ENSG00000111837.12 | DGEA |
| PAPPA2 | ENSG00000116183.11 | DGEA |
| RPE65 | ENSG00000116745.7 | DGEA |
| NUP85 | ENSG00000125450.11 | DGEA |
| TGM3 | ENSG00000125780.12 | DGEA |
| SGCE | ENSG00000127990.19 | DGEA |
| FCHO1 | ENSG00000130475.14 | DGEA |
| DIAPH1 | ENSG00000131504.17 | DGEA |
| PDHA1 | ENSG00000131828.14 | DGEA |
| CRB1 | ENSG00000134376.17 | DGEA |
| TRAFD1 | ENSG00000135148.12 | DGEA |
| TSPAN31 | ENSG00000135452.10 | DGEA |
| SCEL | ENSG00000136155.17 | DGEA |
| ANXA7 | ENSG00000138279.16 | DGEA |
| TBCK | ENSG00000145348.17 | DGEA |
| PTGES | ENSG00000148344.11 | DGEA |
| CNNM2 | ENSG00000148842.18 | DGEA |
| ADRA2A | ENSG00000150594.7 | DGEA |
| CLIC2 | ENSG00000155962.13 | DGEA |
| PAXIP1 | ENSG00000157212.19 | DGEA |
| LCE2B | ENSG00000159455.9 | DGEA |
| SERPINA12 | ENSG00000165953.10 | DGEA |
| KLK4 | ENSG00000167749.11 | DGEA |
| KLK5 | ENSG00000167754.13 | DGEA |
| CTRB1 | ENSG00000168925.12 | DGEA |
| GP2 | ENSG00000169347.17 | DGEA |
| KLF13 | ENSG00000169926.11 | DGEA |
| NHLH1 | ENSG00000171786.6 | DGEA |
| PNLIP | ENSG00000175535.6 | DGEA |
| MEX3C | ENSG00000176624.12 | DGEA |
| TH | ENSG00000180176.15 | DGEA |
| SPRR4 | ENSG00000184148.4 | DGEA |

| | | |
|---|---|---|
| POU3F2 | ENSG00000184486.10 | DGEA |
| ZNF74 | ENSG00000185252.19 | DGEA |
| MIR22HG | ENSG00000186594.14 | DGEA |
| SERPINA3 | ENSG00000196136.18 | DGEA |
| CAPN8 | ENSG00000203697.12 | DGEA |
| C6orf15 | ENSG00000204542.3 | DGEA |
| PHGR1 | ENSG00000233041.8 | DGEA |
| RP11-155G14.6 | ENSG00000240758.2 | DGEA |
| MYZAP | ENSG00000263155.6 | DGEA |
| PRSS2 | ENSG00000275896.6 | DGEA |
| HHATL | ENSG00000010282.15 | DGEA |
| MYO16 | ENSG00000041515.16 | DGEA |
| SDCCAG8 | ENSG00000054282.16 | DGEA |
| CTDP1 | ENSG00000060069.18 | DGEA |
| COL11A1 | ENSG00000060718.22 | DGEA |
| EVI5 | ENSG00000067208.15 | DGEA |
| GAL | ENSG00000069482.7 | DGEA |
| RPH3A | ENSG00000089169.15 | DGEA |
| CHGB | ENSG00000089199.10 | DGEA |
| DDX24 | ENSG00000089737.17 | DGEA |
| MRPS33 | ENSG00000090263.16 | DGEA |
| CPA1 | ENSG00000091704.10 | DGEA |
| KRT31 | ENSG00000094796.5 | DGEA |
| CHGA | ENSG00000100604.13 | DGEA |
| APMAP | ENSG00000101474.12 | DGEA |
| CAB39L | ENSG00000102547.19 | DGEA |
| CNFN | ENSG00000105427.10 | DGEA |
| GLIS3 | ENSG00000107249.24 | DGEA |
| ELMOD1 | ENSG00000110675.13 | DGEA |
| BMP5 | ENSG00000112175.8 | DGEA |
| CCDC181 | ENSG00000117477.12 | DGEA |
| SPP1 | ENSG00000118785.15 | DGEA |
| DBH | ENSG00000123454.12 | DGEA |
| WNT1 | ENSG00000125084.12 | DGEA |
| SLURP1 | ENSG00000126233.2 | DGEA |
| KRT34 | ENSG00000131737.7 | DGEA |
| EMILIN2 | ENSG00000132205.11 | DGEA |
| H3-3B | ENSG00000132475.10 | DGEA |
| FADS2 | ENSG00000134824.14 | DGEA |
| ERCC5 | ENSG00000134899.24 | DGEA |
| KRT85 | ENSG00000135443.8 | DGEA |
| IL36RN | ENSG00000136695.15 | DGEA |
| LOXL4 | ENSG00000138131.4 | DGEA |
| CELA3A | ENSG00000142789.20 | DGEA |
| BICD1 | ENSG00000151746.15 | DGEA |
| CPB1 | ENSG00000153002.12 | DGEA |

| | | |
|---|---|---|
| LPCAT1 | ENSG00000153395.10 | DGEA |
| MCU | ENSG00000156026.14 | DGEA |
| DGKI | ENSG00000157680.16 | DGEA |
| UBN2 | ENSG00000157741.15 | DGEA |
| CPA2 | ENSG00000158516.12 | DGEA |
| VWA5B1 | ENSG00000158816.16 | DGEA |
| ACOX1 | ENSG00000161533.12 | DGEA |
| CTRC | ENSG00000162438.12 | DGEA |
| FRZB | ENSG00000162998.5 | DGEA |
| EN2 | ENSG00000164778.4 | DGEA |
| OSR2 | ENSG00000164920.9 | DGEA |
| CTRB2 | ENSG00000168928.13 | DGEA |
| VCX3A | ENSG00000169059.12 | DGEA |
| KRT86 | ENSG00000170442.12 | DGEA |
| CEL | ENSG00000170835.17 | DGEA |
| PPID | ENSG00000171497.5 | DGEA |
| LIPM | ENSG00000173239.14 | DGEA |
| XCR1 | ENSG00000173578.9 | DGEA |
| CST6 | ENSG00000175315.3 | DGEA |
| ALS2CL | ENSG00000178038.17 | DGEA |
| KCTD12 | ENSG00000178695.6 | DGEA |
| FANCB | ENSG00000181544.15 | DGEA |
| EFHC2 | ENSG00000183690.13 | DGEA |
| LPAR5 | ENSG00000184574.10 | DGEA |
| KRTAP9-8 | ENSG00000187272.7 | DGEA |
| KRTAP1-1 | ENSG00000188581.9 | DGEA |
| SLC22A25 | ENSG00000196600.12 | DGEA |
| MRPL21 | ENSG00000197345.13 | DGEA |
| PRB3 | ENSG00000197870.12 | DGEA |
| AADACL2 | ENSG00000197953.6 | DGEA |
| KRTAP9-9 | ENSG00000198083.9 | DGEA |
| KRTAP4-6 | ENSG00000198090.3 | DGEA |
| KRTAP4-1 | ENSG00000198443.7 | DGEA |
| KCNRG | ENSG00000198553.9 | DGEA |
| C1orf68 | ENSG00000198854.5 | DGEA |
| Y_RNA | ENSG00000201913.1 | DGEA |
| HSD3B2 | ENSG00000203859.10 | DGEA |
| KRTAP9-3 | ENSG00000204873.5 | DGEA |
| PRSS1 | ENSG00000204983.14 | DGEA |
| KRT81 | ENSG00000205426.10 | DGEA |
| KRTAP4-9 | ENSG00000212722.7 | DGEA |
| KRTAP3-3 | ENSG00000212899.3 | DGEA |
| KRTAP3-2 | ENSG00000212900.3 | DGEA |
| CELA2B | ENSG00000215704.10 | DGEA |
| CELA3B | ENSG00000219073.8 | DGEA |
| PPP1R3G | ENSG00000219607.4 | DGEA |

| | | |
|---|---|---|
| PRB4 | ENSG00000230657.8 | DGEA |
| LCE6A | ENSG00000235942.3 | DGEA |
| KRTAP9-2 | ENSG00000239886.5 | DGEA |
| KRTAP10-5 | ENSG00000241123.1 | DGEA |
| KRTAP9-4 | ENSG00000241595.2 | DGEA |
| USP17L22 | ENSG00000248933.3 | DGEA |
| PGA5 | ENSG00000256713.8 | DGEA |
| LINC02412 | ENSG00000258171.2 | DGEA |
| CTC-518B2.9 | ENSG00000268906.1 | DGEA |
| RP11-196G11.3 | ENSG00000280160.1 | DGEA |
