## Supplementary Figures for "Gene expression profiling beyond Breslow thickness and ulceration for prediction of distant metastases in early-stage melanoma: the population-based Dutch Early-Stage Melanoma (D-ESMEL) study"

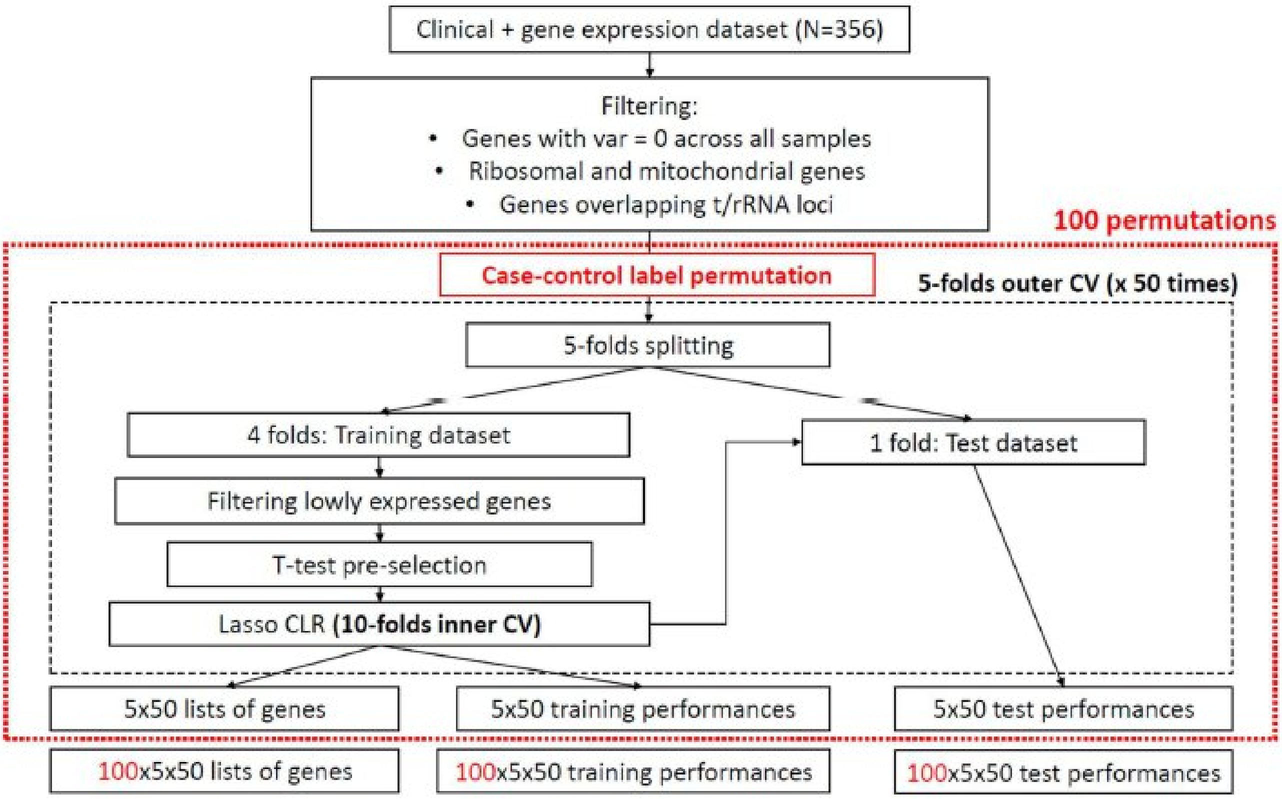


**Supplementary Figure 1.** Schematic of the double-loop cross-validation (DLCV) procedure used for gene selection and model evaluation. After filtering, the dataset (n = 356) was split into 5 outer folds, repeated 50 times. Within each training set, lowly expressed genes were removed, and paired t-tests were used for pre-selection. LASSO-penalized conditional logistic regression with 10-fold inner cross-validation was applied. Model performance was assessed on the outer test folds. To evaluate robustness of selected features, case-control labels were permuted 100 times.

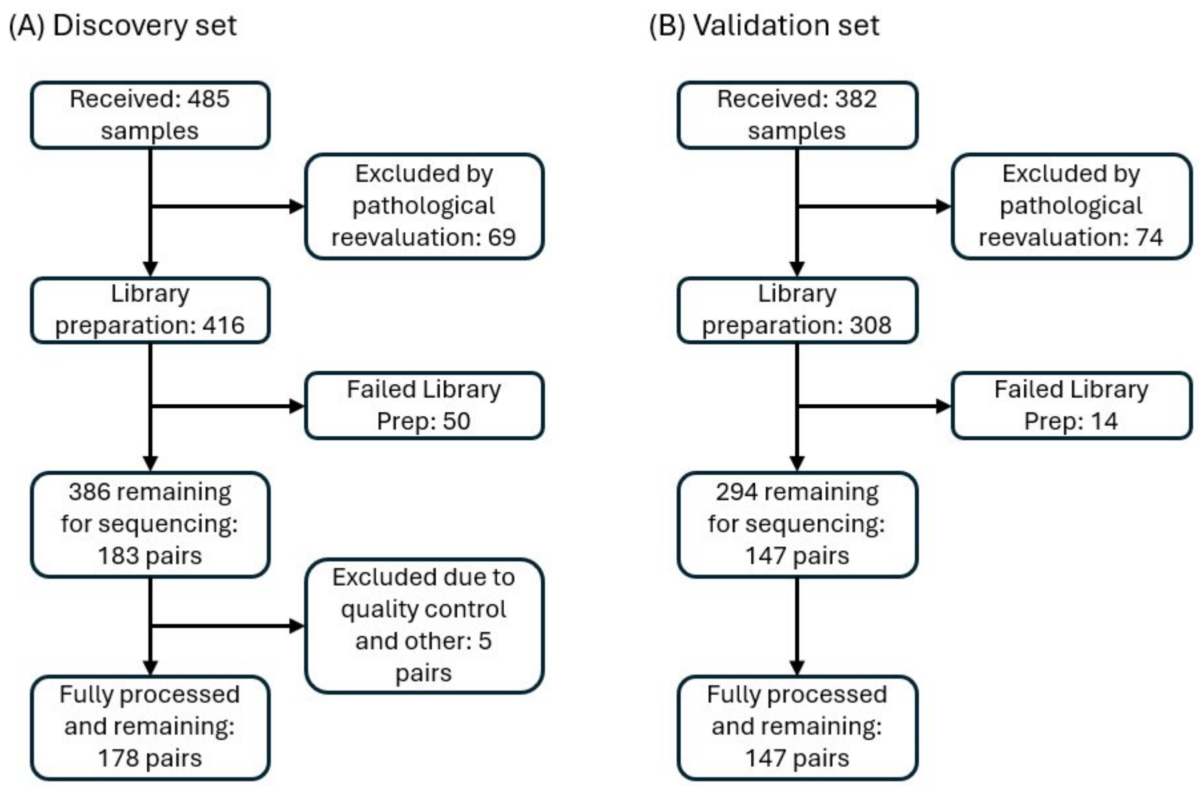


**Supplementary Figure 2**. Flowchart of sample processing and sample exclusion.

A. Discovery set
B. Validation Cohort, model development subset and independent validation subset taken together

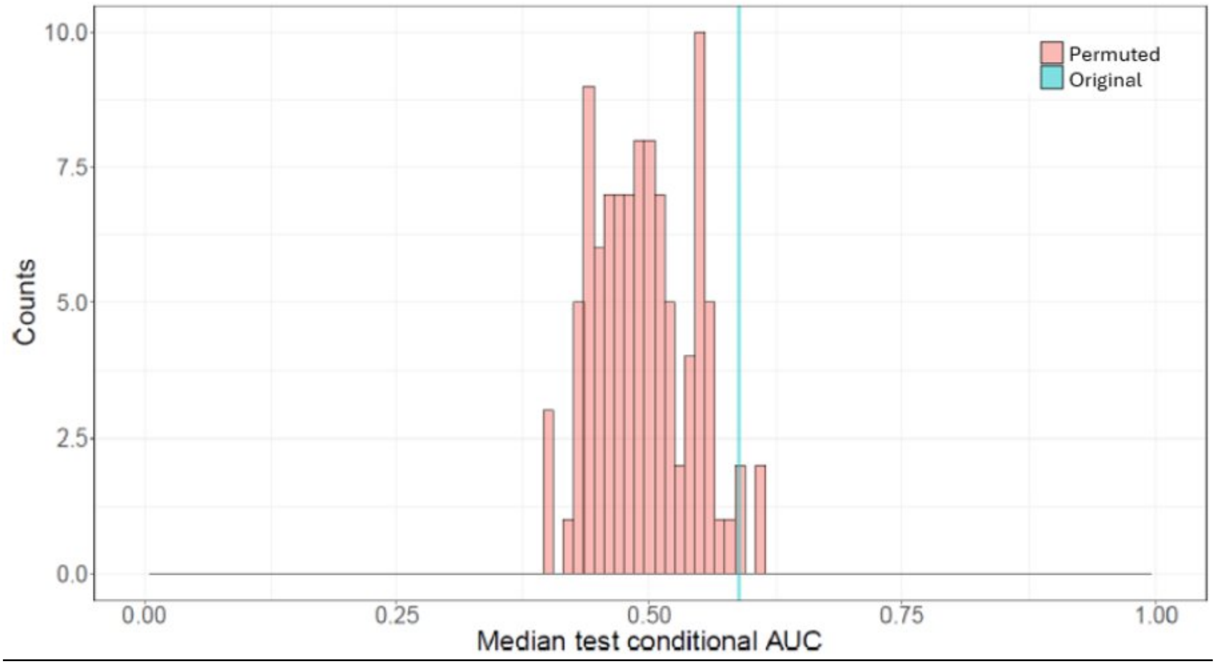

**Supplementary Figure 3**: The performances of models built on permuted and original data are compared as histogram of medians of test conditional AUCs for the 100 permutations and the median test conditional AUC in the original data (light blue line).

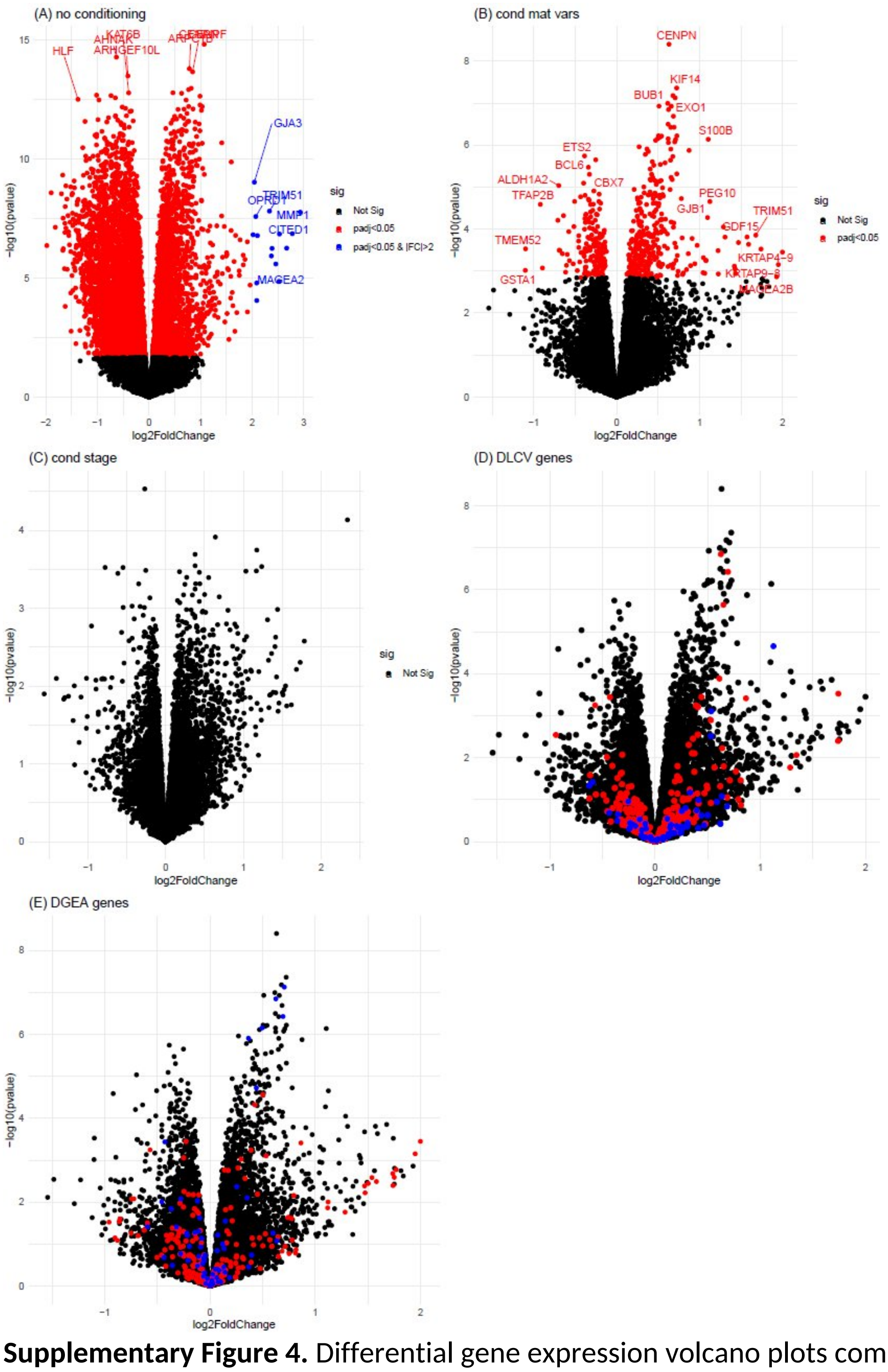


**Supplementary Figure 4.** Differential gene expression volcano plots comparing early-stage melanoma cases versus controls in the model development subset of the validation cohort. (A) Unadjusted analysis.

(B) Conditional analysis accounting for age, sex, Breslow thickness, and ulceration.
(C) Conditional analysis accounting for AJCC stage.
(D) Genes identified with the double-loop cross-validation approach in the discovery set are highlighted in red (*n* = 350); the 64 genes most frequently selected across folds are shown in blue. Conditioned on age, sex, Breslow thickness, and ulceration.
(E) Genes identified with differential gene expression analyses in the discovery set are shown in red (*n* = 288); the 64 most frequently selected genes are shown in blue. Conditioned on age, sex, Breslow thickness, and ulceration.

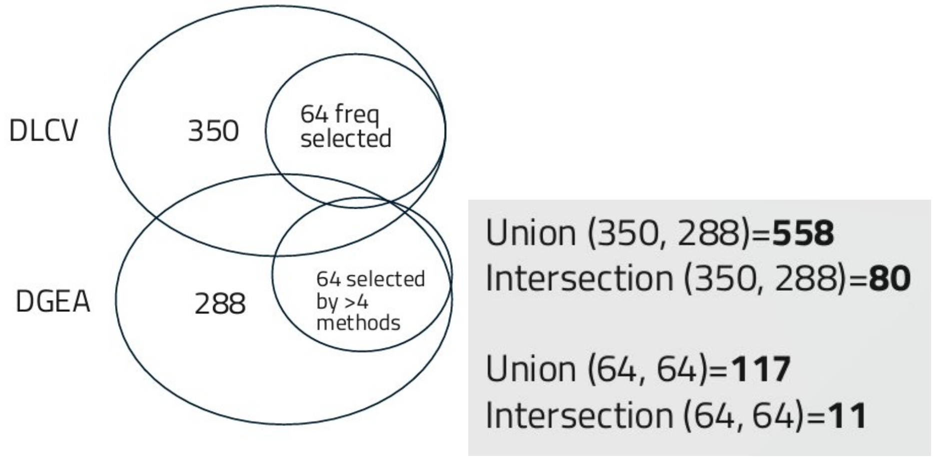


**Supplementary Figure 5**. Selection of subsets of genes with higher selection frequency in the discovery set.

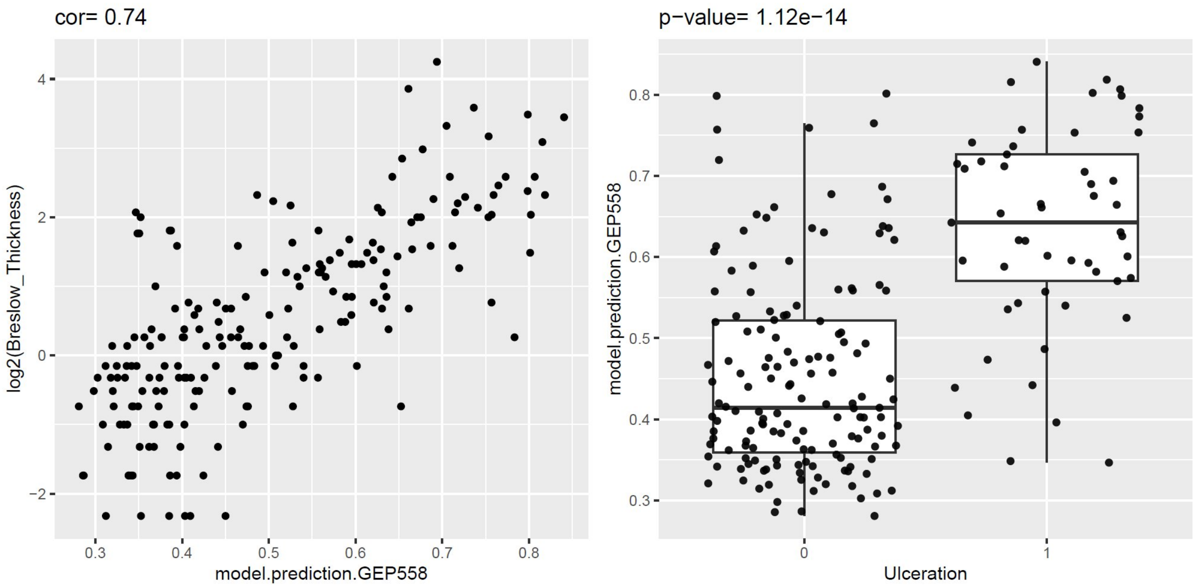


**Supplementary Figure 6.** Association of the 558 discovery genes with Breslow thickness and ulceration in the model development subset of the validation cohort.
A. Correlation between the aggregated prediction score based on the 558 discovery genes (GEP558) and Breslow thickness ($log_2$-transformed), showing a strong positive correlation (r = 0.74).

B. Distribution of prediction scores based on 558 genes by ulceration status, demonstrating significantly higher values in ulcerated compared with non-ulcerated tumors ($p = 1.1 \times 10^{-14}$).

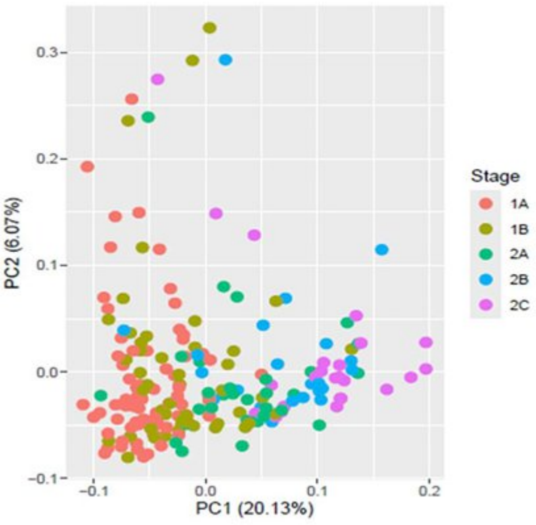


A

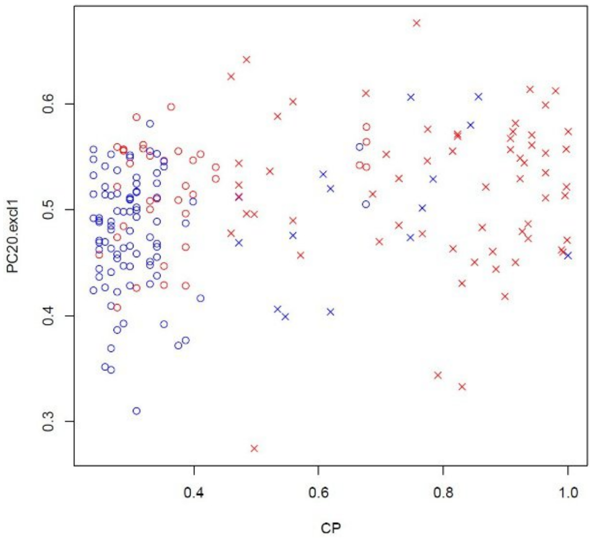


B

**Supplementary Figure 7.** Principal component analysis (PCA) on the 558 discovery genes in the validation cohort.

A. Scatterplot of PC1 versus PC2 colored by AJCC stage.

B Comparison of predicted case probabilities from the clinicopathological (CP) model (logistic regression based on Breslow thickness and ulceration) versus a model based on PCs 2-19 (excluding PC1). PC1 was excluded because it is strongly correlated with the CP score, whereas PCs 2–19 capture components largely independent of CP.

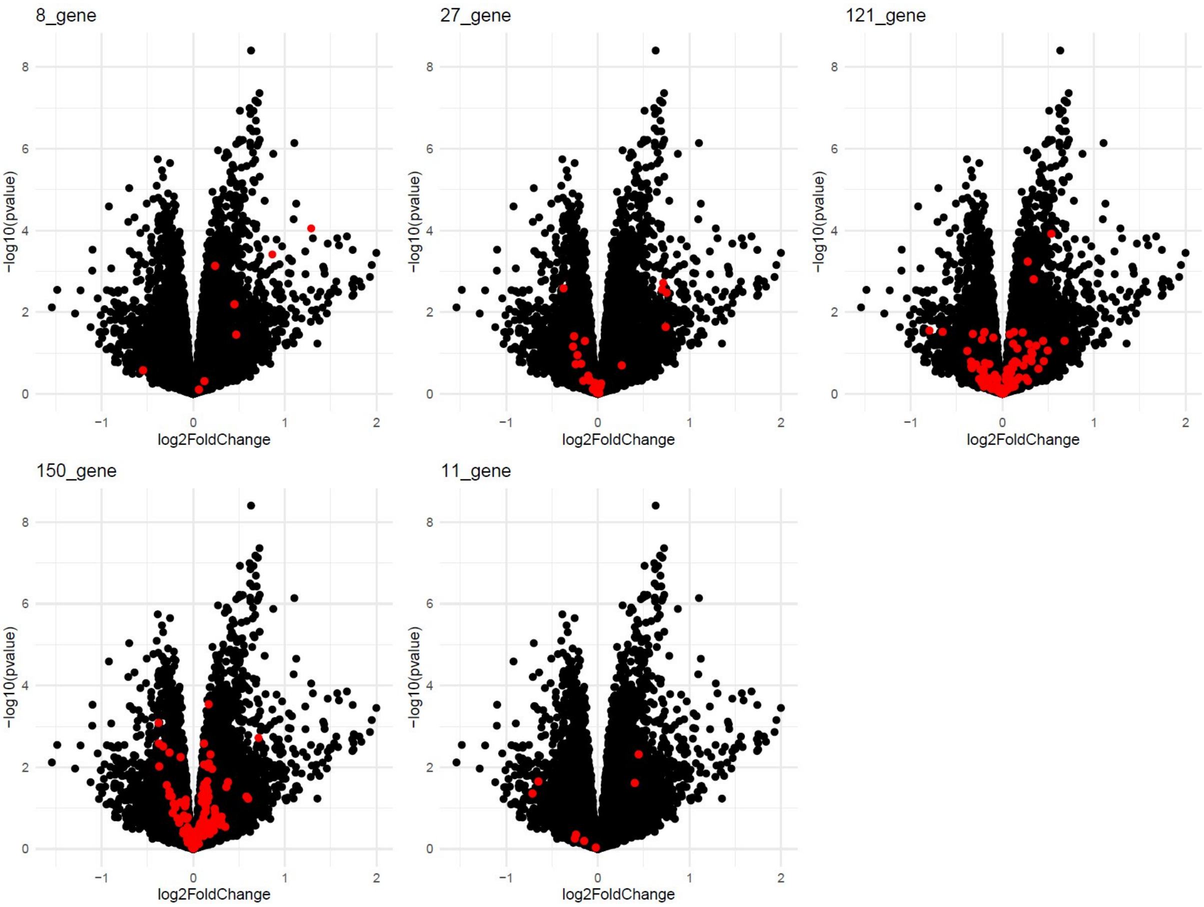


**Supplementary Figure 8.** Differential expression of genes from published gene sets in the validation cohort. Volcano plots show adjusted p-values and effect sizes for external gene sets. Highlighted in red are overlapping genes from our 558 gene list. Conditioned on age, sex, Breslow thickness, and ulceration.
