## Supplementary Methods for "Gene expression profiling beyond Breslow thickness and ulceration for prediction of distant metastases in early-stage melanoma: the population-based Dutch Early-Stage Melanoma (D-ESMEL) study"

### RNA Extraction, Library Preparation, and Sequencing

The collection and processing of formalin-fixed paraffin-embedded (FFPE) primary tumor samples have been previously described (1). Total RNA was extracted using the RNeasy FFPE kit (Qiagen, Hilden, Germany) according to the manufacturer's protocol. RNA concentration and purity were determined using a Nanodrop 2000 spectrophotometer (Thermo Scientific). For library preparation, 50 ng of total RNA was processed using the RNA Prep with Enrichment kit (Illumina, San Diego, CA), incorporating an exome capture approach. The concentration of the generated libraries was measured using a Qubit™ fluorometer (Thermo Fisher Scientific), libraries were pooled and sequencing was performed using the Illumina NextSeq™ system (Illumina, San Diego, CA). The system generated binary base call (BCL) files, which contained base calls for each cycle across all clusters on the flow cell, along with an interop metric file providing sequencing run statistics.

### Bioinformatics Processing and Quality Control

The first step in data processing involved converting BCL files to FASTQ files, followed by demultiplexing using bcl2fastq2 v2.20 (Illumina, San Diego, CA). An in-house analysis pipeline, composed of two separate software packages, Hawkweed and Foxglove, was then applied. Hawkweed generated STAR genome and Salmon transcriptome indices, while Foxglove handled FASTQ file processing, including adapter and quality trimming, read mapping, gene expression quantification, and quality metric extraction.

The Salmon reference transcriptome was constructed from the intersection of the UCSC GoldenPath hg38 reference genome with the Gencode v38 gene model. Sequencing adapters and read segments with a Q-score below 20 were trimmed using TrimGalore. QC metrics from FastQC and GATK were aggregated, and samples identified as outliers or failing thresholds underwent further review.

Principal component analysis (PCA) (stats package v4.2.1; *plotPCA* function in DESeq2 v1.36.0) was used to assess batch effects across library preparations, sequencing runs, and pathology laboratories. When batch effects were detected, the ComBat method was applied for correction.

Read alignment was performed using STAR, generating binary sequence alignment (BAM) files. Read location distribution (coding, intronic, untranslated regions (UTR), intergenic, ribosomal) and transcript coverage normalization were assessed using Picard/GATK CollectRnaSeqMetrics. Gene expression quantification was conducted using Salmon, producing counts and Transcripts-Per-Million (TPM) for each transcript. Gene-level expression values were obtained using the tximport package.

The versions of software used in Hawkweed and Foxglove (v2.04) included STAR (v2.7.10a) (2), Salmon (v1.9.0) (3), FastQC (v0.11.9) (4), TrimGalore (v0.6.6), cutadapt (v3.4), seqtk (v1.3), GATK (4.2.0.0) and samtools/htslib (v1.12) (5) in R version 4.4.0.

#### Details on differential gene expression and modelling

To identify a set of genes that could distinguish cases from controls, we applied two complementary analytical approaches on the discovery set: differential gene expression analysis (DGEA) and discriminative modelling.
DGEA was conducted using multiple statistical frameworks, including DESeq2 (v1.36.0), edgeR (v.3.22), limma (v3.62.2), and direct linear regression (lm from stats v4.4.0) to assess differential expression. Each method was tested with statistical adjustment for the matched design (matching variables (all methods) and case-control pairs (excluding DESeq2, due to unstable dispersion estimates caused by the high number of case-control pairs)). (6) To derive a final set of differentially expressed genes for further model development, a union-based selection approach was applied. From each method, the 100 genes with the smallest *p*-values were selected. Rather than relying on a single statistical model, the final DGEA-based set was determined by taking the union of all selected genes across models.
In parallel, we used conditional logistic regression (CLR) as the basis for discriminative modeling, allowing for internal validation of the selected gene set. Feature selection and parameter estimation were performed using a penalized maximum likelihood estimation algorithm with least absolute shrinkage and selection operator (LASSO) to prevent overfitting and improve model generalization (7, 8).To provide a robust and unbiased assessment of model performance, double-loop cross-validation (DLCV) was implemented. DLCV consisted of an outer loop for model selection and an inner loop for tuning the LASSO penalty. Paired t-tests were used within each training set to rank genes by significance, and the top 5–1000 genes were used as input features to a conditional logistic regression model. The LASSO penalty parameter was optimized via 10-fold inner cross-validation. Model accuracy, as area under the curve (AUC), was evaluated by repeating this process 50 times. To determine whether the observed performance and gene selection were due to chance, case-control labels were permuted 100 times, allowing identification of genes associated with metastases selected with significantly higher frequency. (Supplementary Figure 1)
The statistical analyses were performed in R v4.4.0. A two-sided p-value of 0.05 was considered statistically significant

### References

1. Zhou C, Mooyaart AL, Kerkour T, Louwman MWJ, Wakkee M, Li Y, et al. The Dutch Early-Stage Melanoma (D-ESMEL) study: a discovery set and validation cohort to predict the

absolute risk of distant metastases in stage I/II cutaneous melanoma. Eur J Epidemiol. 2025;40(1):27-42.
2. Dobin A, Davis CA, Schlesinger F, Drenkow J, Zaleski C, Jha S, et al. STAR: ultrafast universal RNA-seq aligner. Bioinformatics. 2013;29(1):15-21.
3. Patro R, Duggal G, Love MI, Irizarry RA, Kingsford C. Salmon provides fast and bias-aware quantification of transcript expression. Nat Methods. 2017;14(4):417-9.
4. Li B, Dewey CN. RSEM: accurate transcript quantification from RNA-Seq data with or without a reference genome. BMC Bioinformatics. 2011;12:323.
5. McKenna A, Hanna M, Banks E, Sivachenko A, Cibulskis K, Kernytsky A, et al. The Genome Analysis Toolkit: a MapReduce framework for analyzing next-generation DNA sequencing data. Genome Res. 2010;20(9):1297-303.
6. Love MI, Huber W, Anders S. Moderated estimation of fold change and dispersion for RNA-seq data with DESeq2. Genome Biol. 2014;15(12):550.
7. Tibshirani R. Regression selection and shrinkage via the lasso. Journal of the Royal Statistical Society Series B. 1996;58(1):267-88.
8. Reid S, Tibshirani R. Regularization Paths for Conditional Logistic Regression: The clogitL1 Package. J Stat Softw. 2014;58(12).
